# Increased L-type calcium current causes action potential prolongation in Jervell and Lange-Nielsen syndrome and is a drug target

**DOI:** 10.1101/2025.03.20.25324224

**Authors:** Yuko Wada, Marcia A. Blair, Teresa L. Strickland, Julie A. Laudeman, Kyungsoo Kim, M. Lorena Harvey, Joseph F. Solus, Darlene F. Fountain, Bjorn C. Knollmann, M. Benjamin Shoemaker, Prince J. Kannankeril, Dan M. Roden

## Abstract

**Background:** *KCNQ1* loss of function variants are thought to cause type 1 long QT syndrome by reducing *I*_Ks_. However, we have recently reported that pharmacologic block of *I*_Ks_ in human induced pluripotent stem cell-derived cardiomyocytes (iPSC-CMs) produced minimal increases in action potential duration at 90% repolarization (APD_90_), while genetic loss of *KCNQ1* markedly prolonged APD_90_. We sought here to define mechanisms underlying APD prolongation by genetic loss of *KCNQ1*.

**Methods:** We studied iPSC-CMs from population controls, an isogenic *KCNQ1* knock out (KO) line created by a homozygous edit for the R518X loss of function variant, and 2 unrelated patients with the Jervell and Lange-Nielsen syndrome (JLN) due to compound heterozygosity for loss of function *KCNQ1* variants.

**Results:** In both JLN and the KCNQ1-KO lines, *I*_Ks_ was absent, APD_90_ was markedly prolonged, and L-type Ca channel (LTCC) current (*I*_Ca-L_) was significantly increased, 2-3-fold, compared to the control cells with no change in kinetics or gating. RNA-sequencing identified 298 and 584 genes that were up– and down-regulated, respectively, by KCNQ1-KO compared to the isogenic control cells. Gene ontology analysis identified down-regulation of 6 Ca^2+^ channel negative regulatory genes (p=0.0002, FDR=0.02), and in knockdown experiments in wild-type iPSC-CMs, three of these, *CBARP*, *FKBP1B*, and *RRAD*, increased *I*_Ca-L_, and *RRAD* increased APD_90_. A therapeutic low concentration (1 μM) of the Ca channel antagonist diltiazem significantly shortened APD_90_ in the two JLN cell lines and in KCNQ1-KO cells. A single low dose of intravenous diltiazem in one of the JLN patients shortened QTc.

**Conclusions:** These data further support the concept that delayed repolarization in JLN cannot be explained solely by loss of *I*_Ks_. Our findings demonstrate that *KCNQ1* mutations lead to down-regulation of Ca^2+^ channel inhibitory genes, with resultant increased *I*_Ca-L_ that underlies delayed repolarization in JLN. We further propose that diltiazem can be repurposed for treatment of patients with JLN.

## Introduction

Ca^2+^ influx through the L-type calcium channel (LTCC) maintains membrane depolarization and initiates excitation-contraction coupling in cardiomyocytes. Both increased depolarizing current (through LTCC or sodium channels) or decreased repolarizing current (through potassium channels) can lead to prolongation of repolarization, as observed in the congenital long QT syndromes (cLQTS). Gain of function of the depolarizing sodium channel Na_V_1.5 (encoded by *SCN5A*, generating *I*_Na_) and LTCC (encoded by *CACNA1C*, generating *I*_Ca-L_) cause LQT3 and LQT8, respectively, while loss of function of the repolarizing cardiac potassium channels K_V_7.1 (encoded by *KCNQ1*, generating *I*_Ks_) and hERG/K_V_11.1 (generating *I*_Kr_) cause LQT1 and LQT2, respectively. In cLQTS, prolonged repolarization of the cardiac action potential can lead to calcium re-influx through LTCC causing early afterdepolarizations that trigger the life-threatening arrhythmia torsades de pointes (TdP).^1,2^ Patients homozygous or compound heterozygous for *KCNQ1* loss of function variants have a particularly severe form of cLQTS, the Jervell and Lange-Neilsen (JLN) syndrome.

Most TdP events in patients with cLQTS occur during β-adrenergic stimulation, and β-blockers are the primary treatment to reduce the risk of TdP.^3,4^ A widely-accepted framework for understanding the efficacy of β-blockade in LQT1 postulates that adrenergic stimulation increases *I*_Ca-L_ through protein kinase A (PKA) stimulation, which would prolong repolarization, and this effect is blunted or blocked by increases in *I*_Ks_;^5^ loss of function variants in *KCNQ1* therefore result in unopposed repolarization prolongation by increased *I*_Ca-L_ (and attendant arrhythmias) with adrenergic stimulation.

Our recent studies in cardiomyocytes developed from induced pluripotent stem cells (iPSC-CMs) challenge this view.^6^ We have shown that *I*_Ks_ is small in these cells, and that its pharmacological block has a negligible effect to prolong repolarization even during PKA activation; however, genetic ablation of *KCNQ1* (by siRNAs or by loss of function variants) markedly prolonged action potential duration at 90% repolarization (APD_90_), an *in vitro* correlate of QT, indicating that *KCNQ1* has actions beyond encoding Kv7.1 channels.

Here we report that increased APD_90_ in multiple iPSC-CM models of JLN is attributable to increased *I*_Ca-L_, arising from downregulated expression of LTCC regulatory proteins. Low doses of the widely-used calcium channel blocker diltiazem normalized APD_90_ in iPSC-CMs and shortened QTc in a JLN patient. These findings not only reshape our mechanistic understanding of JLN but also suggest LTCC inhibition, with a clinically available drug, to treat JLN.

## Methods

Detailed study methods are presented in Supplemental Methods. For generation of iPSCs, a healthy volunteer and patients gave written informed consent prior to inclusion in the study under IRB approval (#9047, #90544) in accordance with the Declaration of Helsinki. All drugs tested *in vitro* in this study are commercially available and listed in Supplemental Table S1. Key *in vitro* recordings were obtained from at least two independent differentiation batches to minimize batch to batch variability, and data are presented as n/N, where n and N indicate the number of recordings and the number of independent differentiation batches, respectively. The clinical test of diltiazem in patient JLN2 was conducted under IRB approval (#240535) and registered at ClinicalTrials.gov (NCT06534671).

## Ethics & Inclusion statement

Patients with JLN were enrolled in the study after they gave written consent. We enrolled all (two) patients alive at Vanderbilt University Medical Center, regardless of their age, gender, race, or ethnicity.

## Statistics

All data are expressed as mean±S.E. unless otherwise indicated. For continuous variables, the Mann-Whitney U test or Kruskal-Wallis test was employed. Two-tailed p<0.05 was considered as significant. Prism 5.0 (GraphPad Software) and JMP9.0 were used for analysis and illustration generation.

## Data availability

The data that support the findings of this study are available from the corresponding author upon reasonable request.

## Results

### Cases of JLN patients with genetic ablation of *KCNQ1*

We studied the differential contribution of ion currents caused by genetic ablation of *KCNQ1* in three iPSC-CM lines in addition to a population control line (“control”): two iPSC lines from unrelated patients with JLN (here termed JLN1 and JLN2, described further below) and one, isogenic to the control iPSCs, in which *KCNQ1* was knocked out by editing in the loss of function variant R518X (KCNQ1-KO) in both alleles.

The first case (JLN1) carries compound heterozygous variants of *KCNQ1*: a paternally inherited nonsense variant R518X and a *de novo* polyA insertion in exon 15 of the maternal allele. The case presentation of the patient has been detailed previously.^7^ RNA sequencing (RNA-seq) in JLN1 iPSC-CMs identified exon 15 skipping and a resultant frame-shift. Prolonged APD_90_ and the absence of *I*_Ks_ even with acute protein kinase A (PKA) stimulation (forskolin and IBMX) have been reported previously.^6^

The second case (JLN2) carries compound heterozygous variants of *KCNQ1*: a maternally inherited R518X and a paternally inherited splice site variant c.921+1 G>T. The case presentation has been previously reported, as a proband in a family named “JLN-10”.^8^ The splice site variant c.921+1 G>T was not detected in that initial report. The parents were heterozygous for the individual variants and had normal QT intervals. Reverse transcription PCR (RT-PCR) using RNA from JLN2 iPSC-CMs revealed that the splice site variant c.921+1G>T resulted in in-frame exon 6 skipping (Supplemental Figure S1).

The KCNQ1-KO line was created using CRISPR/Cas9 genome-editing to introduce the homozygous R518X variant in the control iPSCs (Supplemental Figure S2). We did not have access to a patient with JLN caused by the homozygous R518X; however, cases of JLN homozygous for R518X have been previously reported from a large JLN cohort.^9^

### Effect of loss of KCNQ1 on APD_90_ and *I*_Ks_

In all three iPSC-CMs lines with genetic ablation of *KCNQ1* (JLN1, JLN2, and KCNQ1-KO), APD_90_ was significantly prolonged compared to control cells (Figure 1A-B).

**Figure 1.**
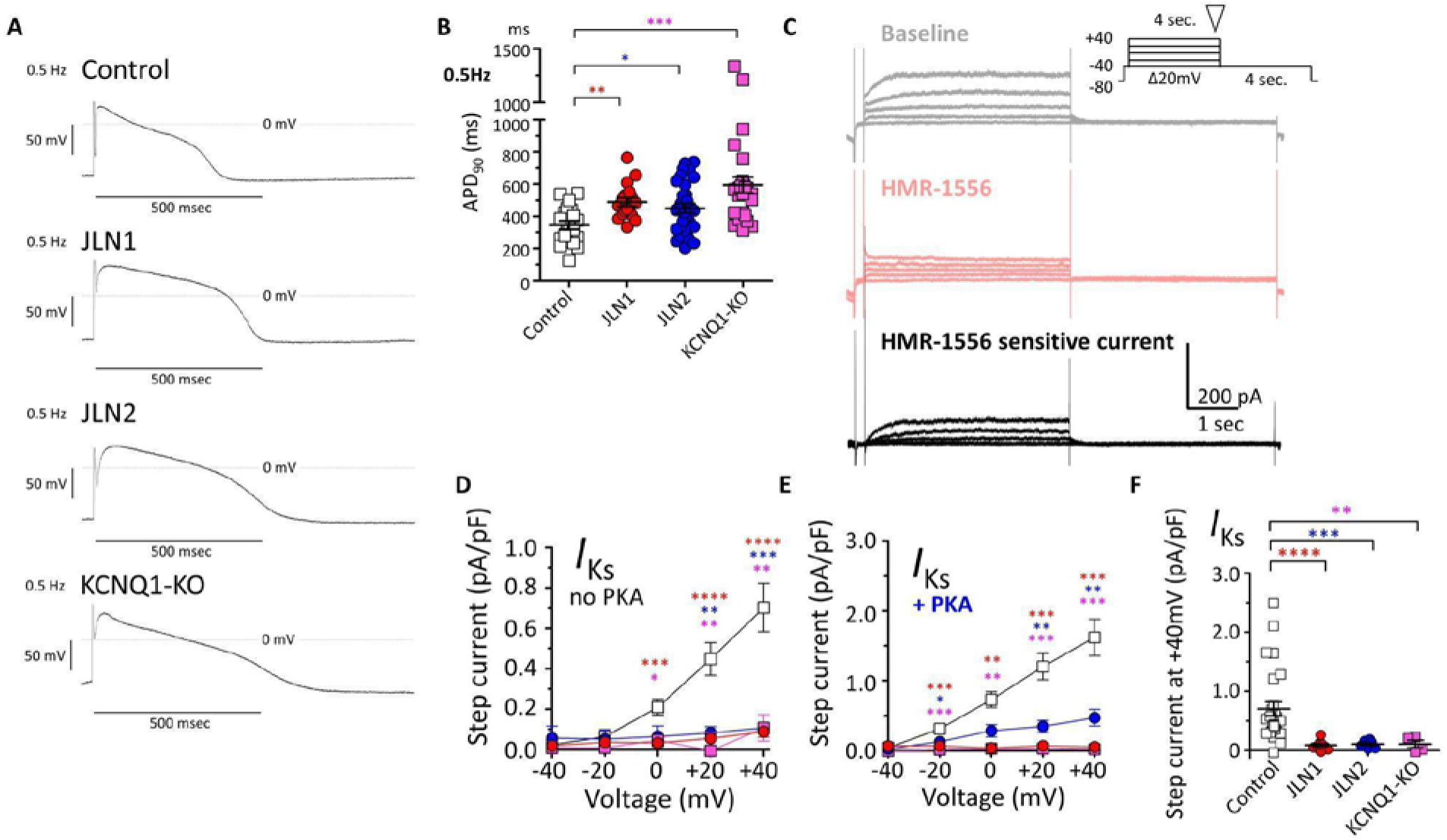
Effect of genetic ablation of *KCNQ1* on action potential duration and *I*_Ks_. **A.** Representative traces of baseline action potential in the cells paced at 0.5 Hz. **B.** Action potential duration at 90% repolarization (APD_90_) in cells paced at 0.5 Hz. Control cells in open squares (n/N=25/5); JLN1 cells in red circles (n/N=17/4); JLN2 cells in blue circles (n/N=31/6); KCNQ1-KO cells in magenta squares (n/N=46/7). **C.** Representative traces of *I*_Ks_ as a current sensitive to the specific blocker HMR-1556. Raw traces before HMR-1556 application are in gray, after HMR-1556 in pink, and after digital subtraction in black. The inset indicates the voltage clamp protocol with an inverted triangle showing where the step current was measured. **D.** *I*_Ks_ current-voltage (*I-V*) relation in the absence of PKA activators. Control cells in open squares (n/N=27/5); JLN1 cells in red circles (n/N=10/5); JLN2 cells in blue circles (n/N=6/2); KCNQ1-KO cells in magenta squares (n/N=4/1). **E.** *I*_Ks_ *I-V* relation with acute exposure to PKA activators (200 μM IBMX + 10 μM forskolin). Control cells in open squares (n/N=10/3); JLN1 cells in red circles (n/N=8/3); JLN2 cells in blue circles (n/N=8/2); KCNQ1-KO cells in magenta squares (n/N=7/2). Note the difference in scale from the data without PKA activators (***D***). **F.** *I*_Ks_ density measured at the end of + 40mV step pulse as shown by the inverted triangle in the inset (***C***). The numbers of cells studied are expressed as n/N, where n indicates number of recordings and N indicates number of differentiation batches. *p<0.05, **p<0.01, ***p<0.001, ****p<0.0001 versus control cells by the Kruskal-Wallis test. Red, blue, and magenta asterisks denote statistics of JLN1, JLN2, and KCNQ1-KO cells against control cells, respectively.

A small *I*_Ks_ was readily recordable in the control cells at baseline (Figure 1C), whereas it was completely absent in JLN1, JLN2, and KCNQ1-KO cells (Figure 1D-F). Acute PKA stimulation significantly increased *I*_Ks_ in the control cells. There was a minimal effect in JLN2 cells and no effect in JLN1 and KCNQ1-KO cells (Figure 1E).

### L-type calcium current was increased in iPSC-CMs with genetic ablation of *KCNQ1*

In all three iPSC-CM lines with genetic ablation of *KCNQ1*, L-type calcium current (*I*_Ca-L_) was markedly increased compared to the control iPSC-CMs (Figure 2A-C). When barium (Ba^2+^) was used as the charge carrier, Ba^2+^ current was increased in all cell lines (as expected); however, the difference in amplitude between control and other lines was markedly attenuated (Figure 2D-F). Voltage-dependence of activation and inactivation were not significantly shifted across all cell lines (Figure 2G). There was also no difference in the fast and slow time constants of inactivation when recorded using Ca^2+^ as the charge carrier (Figure 2H) or Ba^2+^ as the charge carrier (Figure 2I). These results suggest that calcium-dependent inactivation was attenuated in cells with genetic loss of *KCNQ1*.

**Figure 2.**
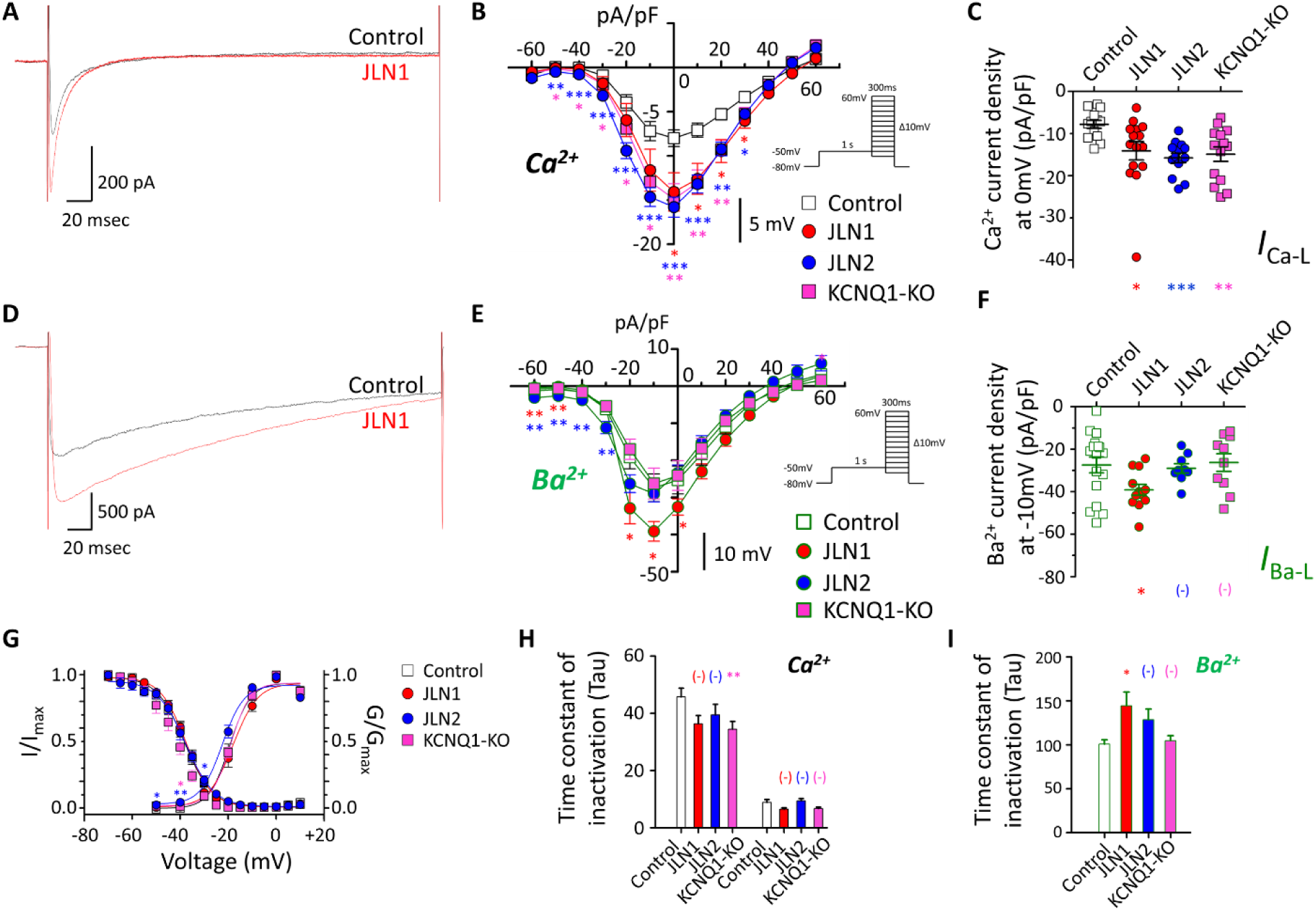
L-type calcium current in iPSC-CMs with genetic ablation of *KCNQ1*. **A.** Raw traces of L-type calcium current (LTCC) at a depolarizing pulse of 0 mV using Ca^2+^ as the charge carrier in the control (black trace) and JLN1 (red trace) cell. **B-C.** The *I-V* relation (***B***) and peak maximal current (at 0 mV, ***C***) for LTCC measured using Ca^2+^ as the charge carrier. Control cells in the open squares (n/N=13/2); JLN1 cells in the red circles (n/N=14/2); JLN2 cells in the blue circles (n/N=14/3); KCNQ1-KO cells in the magenta squares (n/N=14/2). **D.** Raw traces of LTCC at a depolarizing pulse of +10 mV using Ba^2+^ as the charge carrier in the control (black trace) and JLN1 (red trace) cell. **E-F.** The *I-V* relation (***E***) and peak maximal current (at –10 mV, ***F***) for LTCC measured using Ba^2+^ as the charge carrier. Control cells in the open squares (n/N=17/3); JLN1 cells in the red circles (n/N=12/2); JLN2 cells in the blue circle (n/N=9/2); KCNQ1-KO cells in the magenta squares (n/N=10/2). Note the scale difference from (***B)***. **G.** Voltage dependence of activation and inactivation for LTCC using Ca^2+^ as the charge carrier. **H.** Time constants of inactivation measured at 0 mV in Ca^2+^. **I.** Time constant (Tau) of inactivation measured at 0 mV in Ba^2+^. A single time constant was sufficient to describe inactivation in Ba^2+^ (***I***), while 2 were used when Ca^2+^ was the charge carrier (***H***). The numbers of cells studied are expressed as n/N, where n indicates number of recordings and N indicates number of differentiation batches. (-) not significant. *p<0.05, **p<0.01, ***p<0.001 versus control cells by Kruskal-Wallis test. Red, blue, and magenta asterisks denote statistics of JLN1, JLN2, and KCNQ1-KO cells against control cells, respectively.

There were no consistent changes in other key ion currents. In JLN1 and JLN2 cells, *I*_Kr_ was increased (which would shorten APD_90_), while there was no increase in *I*_Kr_ in KCNQ1-KO cells compared to the isogenic control cells (Supplemental Figure S3). There was also no difference in sodium current (*I*_Na_) and sodium-calcium exchanger current (*I*_NCX_) (Supplemental Figure S4A-E).

Taken together, these data demonstrate that genetic loss of *KCNQ1* impaired repolarization through increased Ca^2+^ current.

### Calcium handling was not altered by increased L-type calcium current

We measured diastolic Ca^2+^, calcium transient amplitude, time to baseline (10%, 50%, and 90%), Ca^2+^ content in sarcoplasmic reticulum (SR), fractional Ca^2+^ release from SR, and calcium transient decay rate at room temperature. There were no significant differences in any of these parameters among control and KCNQ1-KO cells (Supplemental Figures S5A-F).

### RNA sequencing identified altered expression of calcium channel regulatory genes

Quantitative PCR did not find altered gene expression of major ion channels (Supplemental Figure S6). To identify pathways and associated ion channel regulatory genes that were affected by the loss of *KCNQ1* and led to the increased availability of LTCC, we sought differentially expressed genes between the isogenic lines (control and KCNQ1-KO). *KCNQ1* itself was most significantly differentially expressed (0.03-fold change, adjusted p=3.9E-94) while there was no difference in expression of other major ion channels, including *CACNA1C*. There were 298 and 584 genes that were significantly up– and down-regulated, respectively (Supplemental Data S1). Gene-ontology analysis identified 154, 24, and 17 pathways that were significantly involved in biological processes (Supplemental Data S2), cellular components (Supplemental Data S3), or molecular functions (Supplemental Data S4). Pathways associated with cardiac depolarization and repolarization are depicted in Supplemental Figures S7-8. These included down-regulation of genes that negatively regulate Ca^2+^ transport (GO1903170; 6 genes [*ATP1A2*, *BIN1*, *CBARP*, *FKBP1B*, *RRAD*, *TGFB1*], p=0.0002, FDR=0.020), and down-regulation of genes involved in Na^+^ transport (GO2000649) and in K^+^ transport (GO:0071805).

### Altered calcium channel regulatory gene expression increased LTCC in iPSC-CMs

To study the effect of the altered calcium channel regulatory genes on LTCC function, we used small interfering RNAs (siRNAs) in the control iPSC-CMs to knock down each of 6 Ca^2+^ transport (GO1903170) genes down-regulated in KCNQ1-KO cells in the RNA-Seq experiments. Knockdown efficiency was assessed by quantitative PCR (Figure 3A), followed by LTCC recordings. Among the 6 genes, 3 (*CBARP*, *FKBP1B*, and *RRAD*) were found to negatively regulate LTCC in iPSC-CMs; knockdown of each of these genes increased *I*_Ca-L_ compared to the negative control iPSC-CMs (Figure 3B-E). Among these three genes, only *RRAD* but not *CBARP* or *FKBP1B* also affected APD_90_; knockdown of *RRAD* prolonged APD_90_ (Figure 3F). Gene-specific knockdown of *KCNQ1* itself significantly increased *I*_Ca-L_ (Figure 3G), replicating our previous finding.^6^

**Figure 3.**
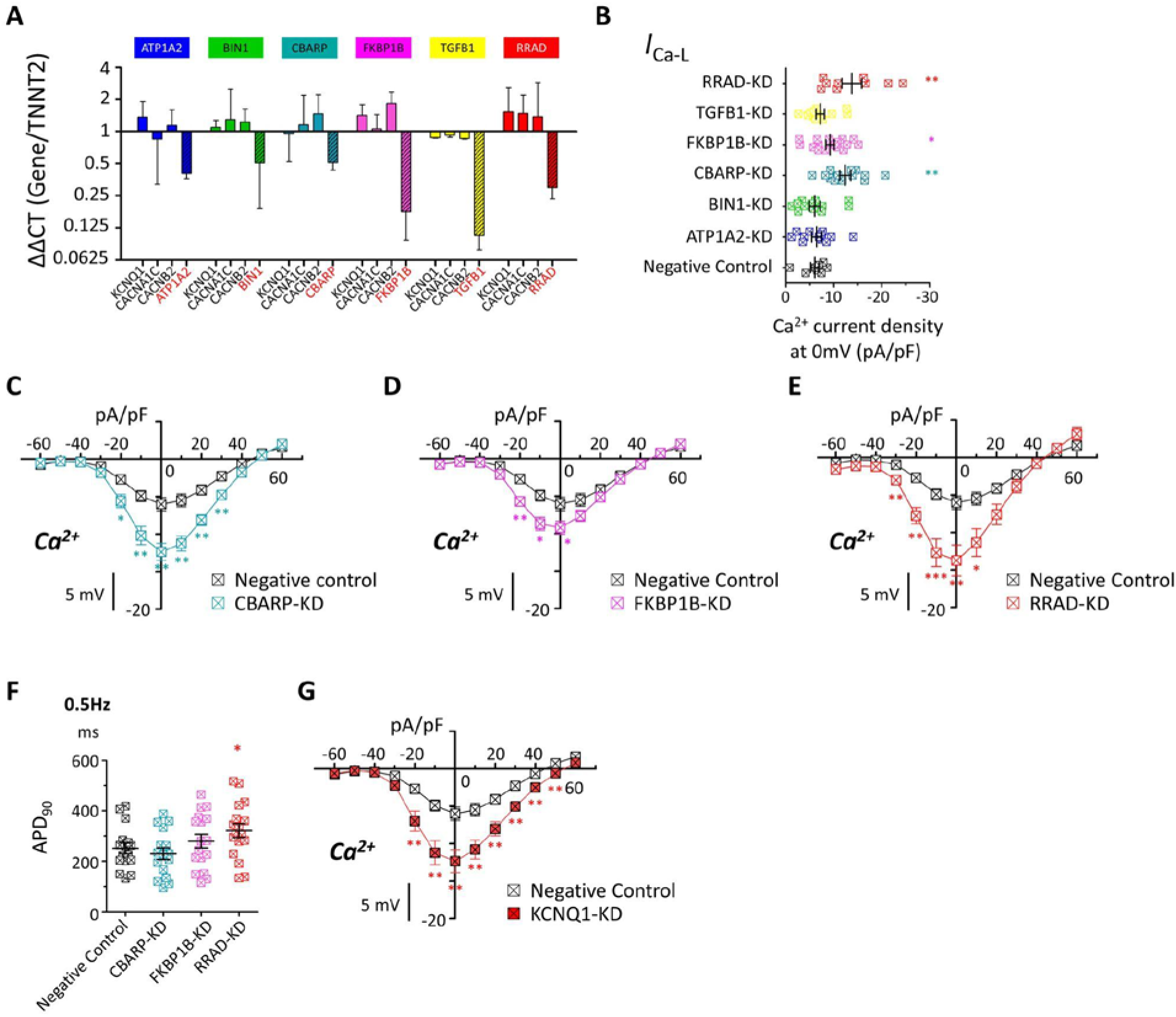
Effects of gene-specific knockdown of calcium channel regulatory genes. **A.** Quantitative PCR showed efficient and specific gene knockdown of the 6 Ca^2+^ regulatory genes studied. Median and interquartile range are shown. N=3-6 per gene target. **B.** The effect of gene-specific knockdown of individual Ca^2+^ regulatory genes on *I*_Ca-L_ measured at 0 mV with Ca^2+^ as the charge carrier: Negative control cells, n/N=8/2; ATP1A2-KD, n/N=12/2; BIN1-KD, n/N=12/2; CBARP-KD, n/N=13/2; FKBP1B-KD, n/N=17/3; TGFB1-KD, n/N=12/2; RRAD-KD, n/N=9/2. **C-E.** The *I-V* relation for LTCC in CBARP (***C***), FKBP1B (***D***), and RRAD (***E***) knockdown cells. **F.** APD_90_ in negative control (n/N=16/3), CBARP (n/N=16/2), FKBP1B (n/N=17/2), and RRAD (n/N=17/2) knockdown cells. **G.** The *I-V* relation for LTCC in *KCNQ1* knockdown cells (n/N=13/2). The numbers of cells studied are expressed as n/N, where n indicates number of recordings and N indicates number of differentiation batches. *p<0.05, **p<0.01, ***p<0.001 versus negative control cells by Mann-Whitney test or Kruskal-Wallis test.

### A low concentration of diltiazem shortened APD in JLN cells

Calcium channel antagonists block LTCC, and some (like verapamil) also block *I*_Kr_. This “balanced block” feature has prompted studies on drug repurposing to blunt prolonged repolarization; in one study, verapamil produced only mild QTc prolongation, and diltiazem had no effect on QTc prolongation induced by the *I*_Kr_ blocker dofetilide.^10^ In control iPSC-CMs, we defined the IC_50_ for *I*_Kr_ block by diltiazem as 24.4 μM and by verapamil 0.80 μM (Supplemental Figure S9); the therapeutic concentrations are 0.6-0.9 μM for both drugs.^10^ In the control cells, diltiazem at up to 1.0 μM minimally affected APD_90_, while significant APD_90_ shortening was seen in all three disease model lines at 0.1-1.0 μM (Figure 4A-C).

**Figure 4.**
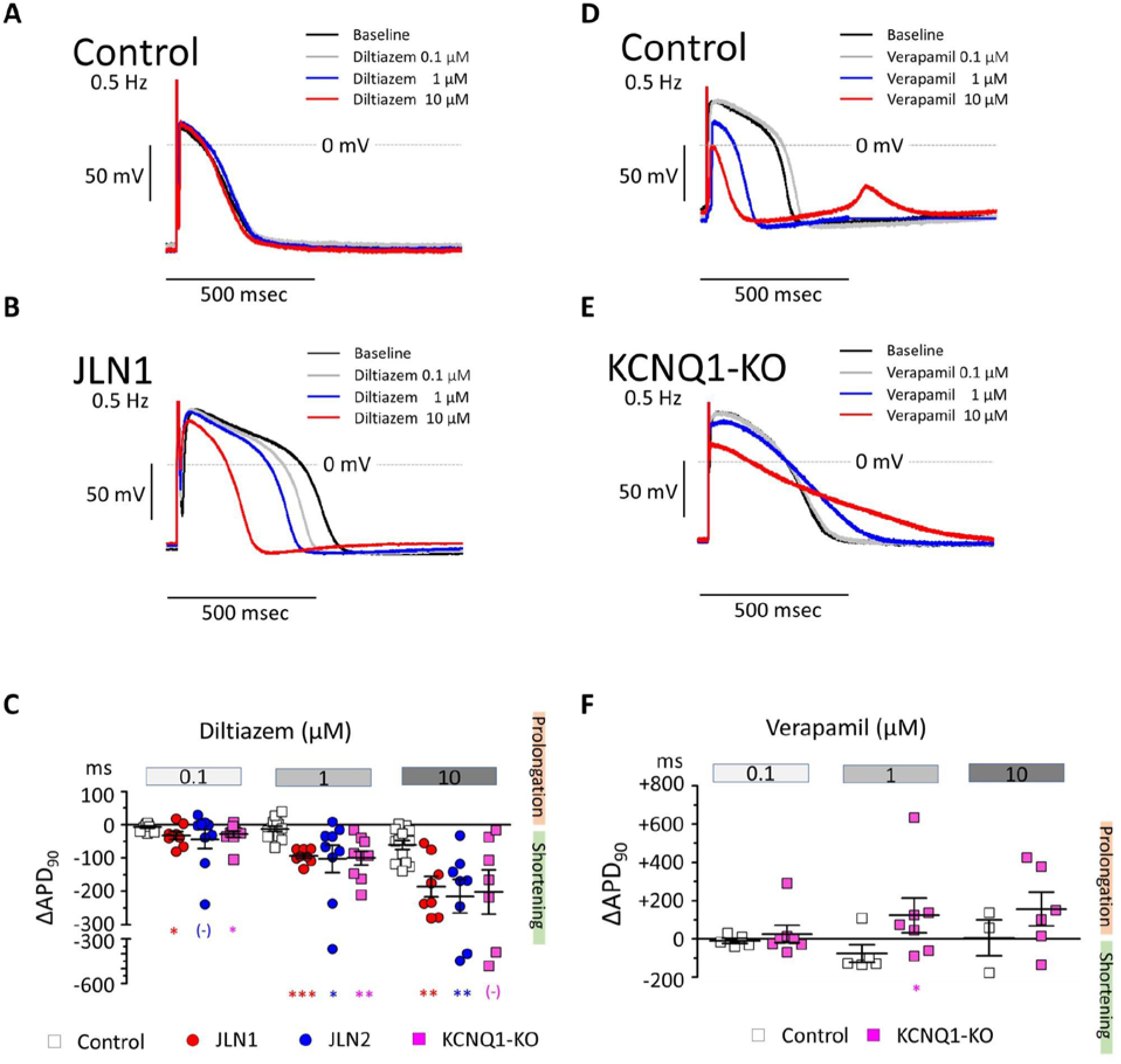
Effect of the Ca channel antagonists diltiazem and verapamil on action potential duration. **A-B.** Representative traces of action potentials in control (***A)*** and JLN1 (***B***) iPSC-CMs during continuous recording with serially increasing concentrations of diltiazem at 0 μM (baseline, black trace), 0.1 μM (gray trace), 1 μM (blue trace), and 10 μM (red trace). **C.** Summary data showing the effect of diltiazem on APD_90_ in the control (open squares, n/N=13/3), JLN1 (red circles, n/N=8/4), JLN2 (blue circle, n/N=9/2), and KCNQ1-KO cells (magenta squares. n/N=10/3). **D-E.** Representative traces of action potentials in control (***D***) and KCNQ1-KO (***E***) iPSC-CMs during continuous recording with serially increasing concentrations of verapamil at 0 μM (baseline, black trace), 0.1 μM (gray trace), 1 μM (blue trace), and 10 μM (red trace). **F.** Summary data showing change in APD_90_ by verapamil in the control (open squares, n/N=5/1) and KCNQ1-KO cells (magenta squares, n/N=7/2). The numbers of cells studied are expressed as n/N, where n indicates number of recordings and N indicates number of differentiation batches. (-) not significant. *p<0.05, **p<0.01, ***p<0.001 versus control cells by Mann-Whitney test or Kruskal-Wallis test. Red, blue, and magenta asterisks denote statistics of JLN1, JLN2, and KCNQ1-KO cells against control cells, respectively, unless otherwise specified.

Verapamil has been reported effective in Timothy syndrome (or LQT8), a rare form of cLQTS caused by a gain-of-function variants in *CACNA1C*.^11^ In the present study, verapamil shortened APD_90_ at 1.0 μM (–76±47 ms) in the control cells (Figure 4D-F). However, by contrast, in the KCNQ1-KO cells, verapamil significantly prolonged APD_90_ at 1.0 μM (+124±91, p=0.03 versus control), consistent with its *I*_Kr_ blocking actions.

### Diltiazem shortened QTc in a patient with JLN

After informed consent, patient JLN2 received a single low dose of intravenous diltiazem (0.25 mg/kg, 21 mg) over 2 minutes while maintained on his usual regimen of oral nadolol (40 mg daily). The QT and rate corrected QT (QTc) were 623/569 msec at baseline (Figure 5A), shortened to 570/546 at end of infusion (2 mins), and showed a peak effect at 5 mins (570/526) (Figure 5B). There was a slight decrease in systolic blood pressure (117/78 mmHg at baseline to 101/62 mmHg at 2 mins) with no significant change in heart rate. QT/QTc returned to baseline (631/558 msec) by 20 minutes. Heart rate, QT, QTc, and blood pressure at baseline and during and post-infusion are shown in Figure 5C.

**Figure 5.**
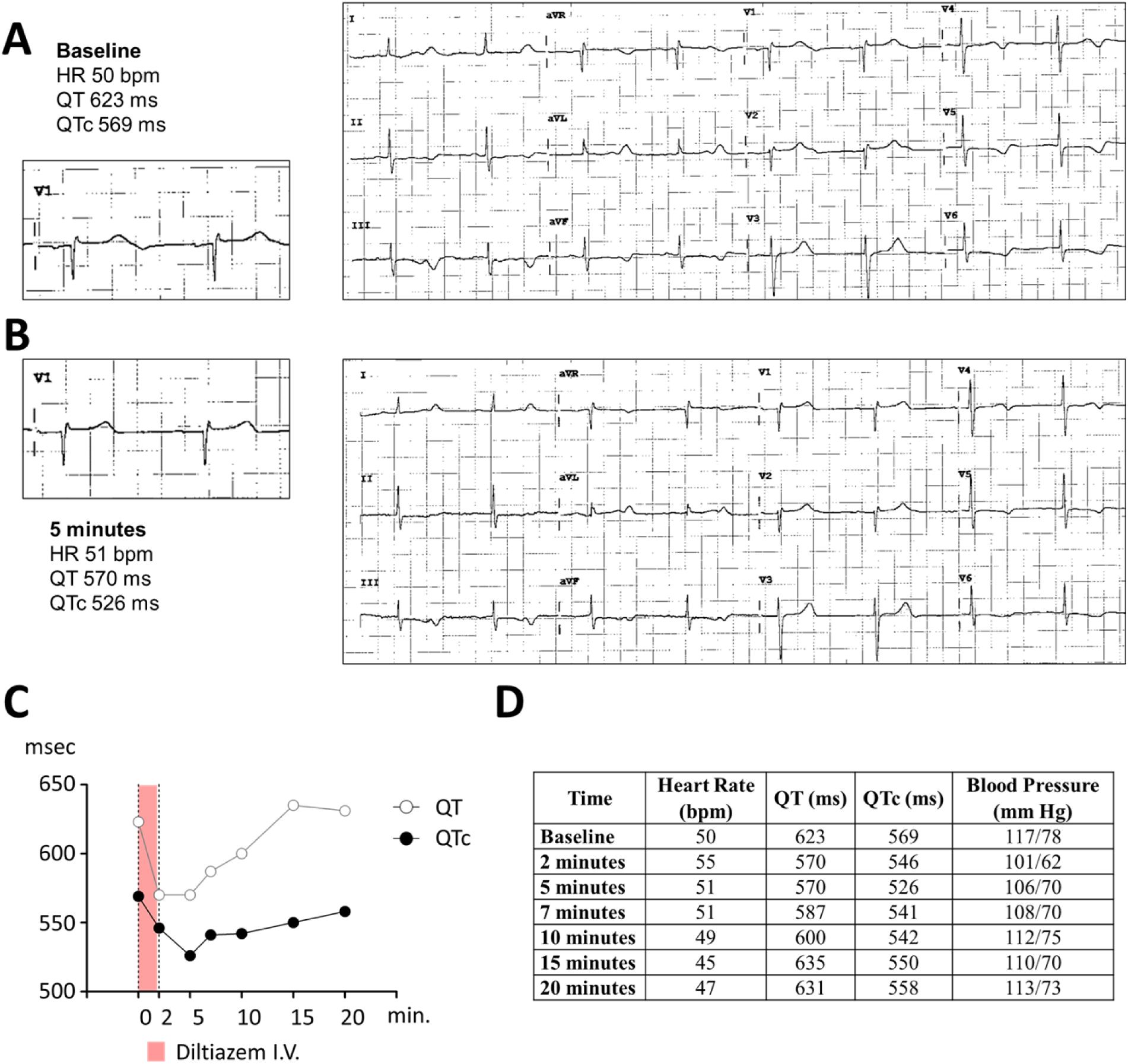
Effect of intravenous diltiazem on QT shortening in a patient with JLN. **A-B.** Electrocardiograms of the patient with JLN (JLN2) at baseline (***A***) and after 5 minutes of diltiazem infusion (***B***). Two consecutive beats in lead V1 are shown in the insets. C. QT and QTc plot during intravenous diltiazem infusion. **D.** Time course of heart rate, QT interval, rate corrected QT (QTc), and blood pressure during and after diltiazem infusion.

## Discussion

### The known direct role of the L-type calcium channel in long QT syndromes

Diagnostic targeted sequencing for symptomatic patients with cLQTS has identified genetic variants in major cardiac ion channels. Molecular genetics and cellular electrophysiology studies have provided insights into the mechanisms whereby altered ion channel function prolongs QT intervals and predisposes to TdP. A direct link between variants in *CACNA1C* and a resultant change in the function of the LTCC has been reported in Timothy syndrome, or LQT8, which is caused by *CACNA1C* gain-of-function variants that disrupt LTCC inactivation. Three auxiliary LTCC subunits (β, γ, and δ) and calmodulin modulate LTCC gating;^12,13^ rare calmodulin variants also cause cLQTS by affecting LTCC inactivation.^14–16^

### The known role of Ca^2+^ controlling *I*_Ks_ associated with long QT syndrome

K_V_7.1 encoded by *KCNQ1* exhibits Ca^2+^-dependent changes in both amplitude and kinetics,^17–19^ and this process is mediated by the direct binding of calmodulin to the carboxy-terminal of K_V_7.1.^20–22^ Both LTCC and K_V_7.1 are highly responsive to β-adrenergic stimulation, and *RRAD* appears to play a role as discussed further below; thus, an imbalance between these inward and outward currents has been thought to account for QT prolongation during exercise leading to TdP in type 1 cLQTS.

### The known role of Ca^2+^ regulatory genes controlling cardiac repolarization

Physiologic and pathophysiologic interactions between calcium signaling and cardiac repolarization have been previously well studied in cardiomyocytes from heart failure models^23–26^ and in simulations of prolonged action potential duration (APD).^26,27^ In these models, prolonged repolarization itself reduces net *I*_Ca-L_, while the time constant of inactivation is prolonged, causing intracellular Ca^2+^ to remain high in failing cardiomyocytes. Although this observation explains why heart failure cardiomyocytes with prolonged APD are susceptible to early and delayed afterdepolarizations, the mechanism by which calcium regulatory pathways are involved and the extent to which LTCCs directly affect QT prolongation in most cLQTS except for LQT8 remains unexamined.

In 2014, a large genome-wide association study probing variability in baseline QT values identified 35 loci.^28^ Of note, these loci included Mendelian cLQTS genes (including *KCNQ1*, *KCNH2*, and *SCN5A*) and further pathway analysis found that three of the top ten pathways were calcium regulatory pathways. These data support the view that calcium regulatory pathways control myocardial repolarization in concert with major cardiac ion channels.

### Studying ion channel interactions beyond a single gene-ion current framework

The gold standard for determining the effect of an ion channel gene variant or a drug on individual currents has been patch-clamping after heterologous expression. Studies showing abnormal *in vitro* function represent evidence for pathogenicity and provide a basis for extrapolating to APD prolongation in cardiomyocytes. However, this extrapolation is predicated on the assumption that variants affect only the target ion current in cardiomyocytes, and this is not always the case. We have previously shown that two ancestrally common variants in the cardiac sodium channel gene *SCN5A* not only destabilize fast inactivation (an effect that would prolong repolarization) but also consistently increase *I*_Kr_ in iPSC-CMs; as a result, repolarization (QT in patients and APD_90_ in cells) is normal but the iPSC-CMs carrying the variants display marked APD_90_ prolongation with exposure to *I*_Kr_ blockers. We have also reported that the contribution of *I*_Ks_ to APD_90_ is nearly negligible even with PKA activation,^6^ forming the basis for the present study.

### What the present study adds

In this study, we implicate dysregulated transcriptional control of calcium current as the primary mechanism underlying QT prolongation when the potassium channel gene *KCNQ1* is knocked out in humans, the JLN syndrome. The R518X homozygous line we generated showed a striking reduction in *KCNQ1* transcripts, and we have also shown that the complex *KCNQ1* mutations seen in the JLN and JLN2 patients also drastically reduce *KCNQ1* transcript abundance. We show here that this loss of *KCNQ1* transcripts is accompanied by a marked increase in *I*_Ca-L_ and this accounts for the prolonged APD_90_. Our data demonstrate that the increased LTCC function results from altered expression of calcium channel regulatory genes, through mechanisms that remain to be defined. We identified three genes, including *RRAD*, as being down-regulated by *KCNQ1* knockout and increasing *I*_Ca-L_ with knockdown in control iPSC-CMs, consistent with previous reports^29–31^ A number of studies have implicated *RRAD* as a key mediator of β-adrenergic augmentation of cardiac contractility, but with no effect on repolarization in mouse knockout models;^31–33^ by contrast, *RRAD* knockdown prolonged APD_90_ in the control human iPSC-CMs. Knockdown of two other genes, *CBARP* and *FKBP1B*, also increased *I*_Ca-L_, but neither altered APD_90_. These results are consistent with the view that *KCNQ1* serves as a network hub controlling repolarization through the combined expression of multiple Ca^2+^ regulatory genes in the human heart.

Building on these results, we showed that diltiazem shortened APD_90_ at a therapeutic low concentration in all lines with genetic ablation of *KCNQ1* but not in the control cells. Unlike diltiazem, verapamil (which has been tested in cLQTS^34^) blocked *I*_Kr_ at near therapeutic concentrations and prolonged APD_90_ in the KCNQ1-KO cells. Importantly, diltiazem was FDA approved for medical use in 1982 and is very widely used in blood pressure and supraventricular arrhythmia management. We tested diltiazem in the patient (JLN2) and observed an immediate shortening of the QT interval.

### Limitations

The molecular links between calcium channel regulatory gene products and the loss of *KCNQ1* remain to be completely defined. The additive effects of diltiazem and β-blockers on APD_90_ in iPSC-CMs need to be addressed. Further studies will be required to establish the extent of QT shortening with chronic oral therapy. The extent to which these findings apply to the commoner autosomal dominant type 1 long QT syndrome is unknown.

### Identification of druggable genes within complex ion channel interactions

Since the mid-1990s, genetic screening for long QT syndrome has significantly improved genotype-based treatment decisions.^35,36^ However, the major treatment option for all forms, antedating the genetic era, remains β-blockade. The ability to generate iPSC-CMs now offers the opportunity of studying not only individual components of ion currents but also their interactions and the “compensatory changes” we have now described. This framework has now defined a new target for intervention in JLN, and this approach may aid in further identifying druggable genes in other types of cLQTS and thus provide patients with better treatment options.

## Conclusion

Genetic loss of *KCNQ1* causes severe APD_90_ prolongation in iPSC-CMs from patients with JLN, and we show here that the unexpected underlying mechanism is increased LTCC activity resulting from altered expression of calcium regulatory genes. The Ca channel antagonist diltiazem shortened APD_90_ in JLN iPSC-CMs at a therapeutic low concentration. Diltiazem effectively shortened QT in the patient with JLN whose iPSC-CMs showed high sensitivity to diltiazem. These data support further studies of diltiazem in patients with JLN.

## Acknowledgement

Flow Cytometry experiments were performed in the Vanderbilt Flow Cytometry Shared Resource supported by the Vanderbilt Ingram Cancer Center (P30 CA068485) and the Vanderbilt Digestive Disease Research Center (DK058404). RNA sequencing and data analysis was performed in the Vanderbilt Technologies for Advanced Genomics (VANTAGE) and Vanderbilt Technologies for Advanced Genomics Analysis and Research Design (VANGARD), respectively, supported in part by the Vanderbilt Vision Center (P30 EY08126).

## Funding

This project was supported by National Institute of Health (R01 HL164675 to D.M.R. and UL1TR002243), the American Heart Association (23CDA1048873 to Y.W.).

## Disclosures

All authors report no conflict of interest.

## Notes

### Competing Interest Statement

The authors have declared no competing interest.

### Author Declarations

Ethics committee/IRB of Vanderbilt University Medical Center gave ethical approval for this work under the IRB numbers 9047, 90544, and 240535.

## References

1. Roden DM, Anderson ME. The pause that refreshes, or does it? Mechanisms in torsades de pointes. Heart. 2000;84:235–237. PMID:10956280.

2. Landstrom AP, Dobrev D, Wehrens XHT. Calcium Signaling and Cardiac Arrhythmias. Circ Res. 2017;120:1969–1993. PMID:28596175. PMC5607780

3. Schwartz PJ, Locati E. The idiopathic long QT syndrome: pathogenetic mechanisms and therapy. Eur Heart J. 1985;6 Suppl D:103–114. PMID:2867907.

4. Moss AJ, Schwartz PJ, Crampton RS, Locati E, Carleen E. The long QT syndrome: a prospective international study. Circulation. 1985;71:17–21. PMID:2856865.

5. Xie Y, Grandi E, Puglisi JL, Sato D, Bers DM. β-adrenergic stimulation activates early afterdepolarizations transiently via kinetic mismatch of PKA targets. J Mol Cell Cardiol. 2013;58:153–161. PMID:23481579. PMC3628092

6. Wada Y, Wang L, Hall LD, Yang T, Short LL, Solus JF, Glazer AM, Roden DM. The electrophysiologic effects of KCNQ1 extend beyond expression of IKs: evidence from genetic and pharmacologic block. Cardiovasc Res. 2024;120:735–744. PMID:38442735. PMC11135641

7. Bersell K, Montgomery JA, Kanagasundram AN, Campbell CM, Chung WK, Macaya D, Konecki D, Venter E, Shoemaker MB, Roden DM. Partial Duplication and Poly(A) Insertion in KCNQ1 Not Detected by Next-Generation Sequencing in Jervell and Lange–Nielsen Syndrome. Circ Arrhythm Electrophysiol. 2016;9:e004081.

8. Wei J, Fish FA, Myerburg RJ, Roden DM, George AL Jr. Novel KCNQ1 mutations associated with recessive and dominant congenital long QT syndromes: evidence for variable hearing phenotype associated with R518X. Hum Mutat. 2000;15:387–388. PMID:10737999.

9. Winbo A, Stattin E-L, Nordin C, Diamant U-B, Persson J, Jensen SM, Rydberg A. Phenotype, origin and estimated prevalence of a common long QT syndrome mutation: a clinical, genealogical and molecular genetics study including Swedish R518X/KCNQ1 families. BMC Cardiovasc Disord. 2014;14:22. PMID:24552659. PMC3942207

10. Vicente J, Zusterzeel R, Johannesen L, Ochoa-Jimenez R, Mason JW, Sanabria C, Kemp S, Sager PT, Patel V, Matta MK, Liu J, Florian J, Garnett C, Stockbridge N, Strauss DG. Assessment of multi-ion channel block in a phase I randomized study design: Results of the CiPA phase I ECG biomarker validation study. Clin Pharmacol Ther. 2019;105:943–953. PMID:30447156. PMC6654598

11. Jacobs A, Knight BP, McDonald KT, Burke MC. Verapamil decreases ventricular tachyarrhythmias in a patient with Timothy syndrome (LQT8). Heart Rhythm. 2006;3:967–970. PMID:16876748.

12. Pérez-García MT, Kamp TJ, Marbán E. Functional properties of cardiac L-type calcium channels transiently expressed in HEK293 cells. Roles of alpha 1 and beta subunits. J Gen Physiol. 1995;105:289–305. PMID:7539049. PMC2216941

13. Morales D, Hermosilla T, Varela D. Calcium-dependent inactivation controls cardiac L-type Ca2+ currents under β-adrenergic stimulation. J Gen Physiol. 2019;151:786–797. PMID:30814137. PMC6571991

14. Crotti L, Johnson CN, Graf E, De Ferrari GM, Cuneo BF, Ovadia M, Papagiannis J, Feldkamp MD, Rathi SG, Kunic JD, Pedrazzini M, Wieland T, Lichtner P, Beckmann B-M, Clark T, Shaffer C, Benson DW, Kääb S, Meitinger T, Strom TM, Chazin WJ, Schwartz PJ, George AL Jr. Calmodulin mutations associated with recurrent cardiac arrest in infants. Circulation. 2013;127:1009–1017. PMID:23388215. PMC3834768

15. Limpitikul WB, Dick IE, Joshi-Mukherjee R, Overgaard MT, George AL Jr, Yue DT. Calmodulin mutations associated with long QT syndrome prevent inactivation of cardiac L-type Ca(2+) currents and promote proarrhythmic behavior in ventricular myocytes. J Mol Cell Cardiol. 2014;74:115–124. PMID:24816216. PMC4262253

16. Rocchetti M, Sala L, Dreizehnter L, Crotti L, Sinnecker D, Mura M, Pane LS, Altomare C, Torre E, Mostacciuolo G, Severi S, Porta A, De Ferrari GM, George AL Jr, Schwartz PJ, Gnecchi M, Moretti A, Zaza A. Elucidating arrhythmogenic mechanisms of long-QT syndrome CALM1-F142L mutation in patient-specific induced pluripotent stem cell-derived cardiomyocytes. Cardiovasc Res. 2017;113:531–541. PMID:28158429.

17. O-Uchi J, Rice JJ, Ruwald MH, Parks XX, Ronzier E, Moss AJ, Zareba W, Lopes CM. Impaired IKs channel activation by Ca(2+)-dependent PKC shows correlation with emotion/arousal-triggered events in LQT1. J Mol Cell Cardiol. 2015;79:203–211. PMID:25479336. PMC4302024

18. Bartos DC, Morotti S, Ginsburg KS, Grandi E, Bers DM. Quantitative analysis of the Ca2+ – dependent regulation of delayed rectifier K+ current IKs in rabbit ventricular myocytes. J Physiol. 2017;595:2253–2268. PMID:28008618. PMC5374113

19. Shugg T, Johnson DE, Shao M, Lai X, Witzmann F, Cummins TR, Rubart-Von-der Lohe M, Hudmon A, Overholser BR. Calcium/calmodulin-dependent protein kinase II regulation of IKs during sustained β-adrenergic receptor stimulation. Heart Rhythm. 2018;15:895–904. PMID:29410121. PMC5984714

20. Ghosh S, Nunziato DA, Pitt GS. KCNQ1 Assembly and Function Is Blocked by Long-QT Syndrome Mutations That Disrupt Interaction With Calmodulin. Circ Res. 2006;98:1048–1054.

21. Shamgar L, Ma L, Schmitt N, Haitin Y, Peretz A, Wiener R, Hirsch J, Pongs O, Attali B. Calmodulin Is Essential for Cardiac IKS Channel Gating and Assembly. Circ Res. 2006;98:1055– 1063.

22. McCormick L, Wadmore K, Milburn A, Gupta N, Morris R, Held M, Prakash O, Carr J, Barrett-Jolley R, Dart C, Helassa N. Long QT syndrome-associated calmodulin variants disrupt the activity of the slow delayed rectifier potassium current. J Physiol. 2023;PMID:37428651.

23. Beuckelmann DJ, Näbauer M, Erdmann E. Intracellular calcium handling in isolated ventricular myocytes from patients with terminal heart failure. Circulation. 1992;85:1046–1055. PMID:1311223.

24. Mukherjee R, Hewett KW, Walker JD, Basler CG, Spinale FG. Changes in L-type calcium channel abundance and function during the transition to pacing-induced congestive heart failure. Cardiovasc Res. 1998;37:432–444. PMID:9614498.

25. Kaprielian R, Wickenden AD, Kassiri Z, Parker TG, Liu PP, Backx PH. Relationship between K+channel down-regulation and [Ca2+]i in rat ventricular myocytes following myocardial infarction. J Physiol. 1999;517 ( Pt 1):229–245. PMID:10226162. PMC2269317

26. Cooper PJ, Soeller C, Cannell MB. Excitation-contraction coupling in human heart failure examined by action potential clamp in rat cardiac myocytes. J Mol Cell Cardiol. 2010;49:911–917. PMID:20430038.

27. Bouchard RA, Clark RB, Giles WR. Effects of action potential duration on excitation-contraction coupling in rat ventricular myocytes. Action potential voltage-clamp measurements: Action potential voltage-clamp measurements. Circ Res. 1995;76:790–801. PMID:7728996.

28. Arking DE, Pulit SL, Crotti L, van der Harst P, Munroe PB, Koopmann TT, Sotoodehnia N, Rossin EJ, Morley M, Wang X, Johnson AD, Lundby A, Gudbjartsson DF, Noseworthy PA, Eijgelsheim M, Bradford Y, Tarasov KV, Dörr M, Müller-Nurasyid M, Lahtinen AM, Nolte IM, Smith AV, Bis JC, Isaacs A, Newhouse SJ, Evans DS, Post WS, Waggott D, Lyytikäinen L-P, Hicks AA, Eisele L, Ellinghaus D, Hayward C, Navarro P, Ulivi S, Tanaka T, Tester DJ, Chatel S, Gustafsson S, Kumari M, Morris RW, Naluai ÅT, Padmanabhan S, Kluttig A, Strohmer B, Panayiotou AG, Torres M, Knoflach M, Hubacek JA, Slowikowski K, Raychaudhuri S, Kumar RD, Harris TB, Launer LJ, Shuldiner AR, Alonso A, Bader JS, Ehret G, Huang H, Kao WHL, Strait JB, Macfarlane PW, Brown M, Caulfield MJ, Samani NJ, Kronenberg F, Willeit J, CARe Consortium, COGENT Consortium, Smith JG, Greiser KH, Meyer Zu Schwabedissen H, Werdan K, Carella M, Zelante L, Heckbert SR, Psaty BM, Rotter JI, Kolcic I, Polašek O, Wright AF, Griffin M, Daly MJ, DCCT/EDIC, Arnar DO, Hólm H, Thorsteinsdottir U, eMERGE Consortium, Denny JC, Roden DM, Zuvich RL, Emilsson V, Plump AS, Larson MG, O’Donnell CJ, Yin X, Bobbo M, D’Adamo AP, et al. Genetic association study of QT interval highlights role for calcium signaling pathways in myocardial repolarization. Nat Genet. 2014;46:826–836. PMID:24952745. PMC4124521

29. Manning JR, Yin G, Kaminski CN, Magyar J, Feng H-Z, Penn J, Sievert G, Thompson K, Jin J-P, Andres DA, Satin J. Rad GTPase deletion increases L-type calcium channel current leading to increased cardiac contraction. J Am Heart Assoc. 2013;2:e000459. PMID:24334906. PMC3886777

30. Ahern BM, Levitan BM, Veeranki S, Shah M, Ali N, Sebastian A, Su W, Gong MC, Li J, Stelzer JE, Andres DA, Satin J. Myocardial-restricted ablation of the GTPase RAD results in a pro-adaptive heart response in mice. J Biol Chem. 2019;294:10913–10927. PMID:31147441. PMC6635439

31. Papa A, Zakharov SI, Katchman AN, Kushner JS, Chen B-X, Yang L, Liu G, Jimenez AS, Eisert RJ, Bradshaw GA, Dun W, Ali SR, Rodriques A, Zhou K, Topkara V, Yang M, Morrow JP, Tsai EJ, Karlin A, Wan E, Kalocsay M, Pitt GS, Colecraft HM, Ben-Johny M, Marx SO. Rad regulation of CaV1.2 channels controls cardiac fight-or-flight response. Nat Cardiovasc Res. 2022;1:1022–1038. PMID:36424916. PMC9681059

32. Papa A, Del Rivero Morfin PJ, Chen B-X, Yang L, Katchman AN, Zakharov SI, Liu G, Bohnen MS, Zheng V, Katz M, Subramaniam S, Hirsch JA, Weiss S, Dascal N, Karlin A, Pitt GS, Colecraft HM, Ben-Johny M, Marx SO. A membrane-associated phosphoswitch in Rad controls adrenergic regulation of cardiac calcium channels. J Clin Invest. 2024;134. PMID:38227371. PMC10904049

33. Katchman AN, Zakharov SI, Bohnen MS, Sanchez Jimenez A, Kushner JS, Yang L, Chen B-X, Nasari A, Liu G, Rabbani DE, Han J, Leu C-S, Pitt GS, Marx SO. Augmented cardiac inotropy by phosphodiesterase inhibition requires phosphorylation of rad and increased calcium current. Circulation. 2024;149:1617–1620. PMID:38739694. PMC11614396

34. Shimizu W, Ohe T, Kurita T, Kawade M, Arakaki Y, Aihara N, Kamakura S, Kamiya T, Shimomura K. Effects of verapamil and propranolol on early afterdepolarizations and ventricular arrhythmias induced by epinephrine in congenital long QT syndrome. J Am Coll Cardiol. 1995;26:1299–1309. PMID:7594047.

35. Younis A, Bos JM, Zareba W, Aktas MK, Wilde AAM, Tabaja C, Bodurian C, Tobert KE, McNitt S, Polonsky B, Shimizu W, Ackerman MJ, Goldenberg I. Association Between Syncope Trigger Type and Risk of Subsequent Life-Threatening Events in Patients With Long QT Syndrome. JAMA Cardiol. 2023;PMID:37436769. PMC10339217

36. Kaizer AM, Winbo A, Clur S-AB, Etheridge SP, Ackerman MJ, Horigome H, Herberg U, Dagradi F, Spazzolini C, Killen SAS, Wacker-Gussmann A, Wilde AAM, Sinkovskaya E, Abuhamad A, Torchio M, Ng C-A, Rydberg A, Schwartz PJ, Cuneo BF. Effects of cohort, genotype, variant, and maternal β-blocker treatment on foetal heart rate predictors of inherited long QT syndrome. Europace. 2023;25. PMID:37975542. PMC10655062

37. Cadar AG, Feaster TK, Bersell KR, Wang L, Hong T, Balsamo JA, Zhang Z, Chun YW, Nam Y-J, Gotthardt M, Knollmann BC, Roden DM, Lim CC, Hong CC. Real-time visualization of titin dynamics reveals extensive reversible photobleaching in human induced pluripotent stem cell-derived cardiomyocytes. Am J Physiol Cell Physiol. 2020;318:C163–C173. PMID:31747312. PMC6985833

38. Bersell KR, Yang T, Mosley JD, Glazer AM, Hale AT, Kryshtal DO, Kim K, Steimle JD, Brown JD, Salem J-E, Campbell CC, Hong CC, Wells QS, Johnson AN, Short L, Blair MA, Behr ER, Petropoulou E, Jamshidi Y, Benson MD, Keyes MJ, Ngo D, Vasan RS, Yang Q, Gerszten RE, Shaffer C, Parikh S, Sheng Q, Kannankeril PJ, Moskowitz IP, York JD, Wang TJ, Knollmann BC, Roden DM. Transcriptional Dysregulation Underlies Both Monogenic Arrhythmia Syndrome and Common Modifiers of Cardiac Repolarization. Circulation. 2023;147:824–840. PMID:36524479. PMC9992308

39. Parikh SS, Blackwell DJ, Gomez-Hurtado N, Frisk M, Wang L, Kim K, Dahl CP, Fiane A, Tønnessen T, Kryshtal DO, Louch WE, Knollmann BC. Thyroid and glucocorticoid hormones promote functional T-tubule development in human-induced pluripotent stem cell-derived cardiomyocytes. Circ Res. 2017;121:1323–1330. PMID:28974554. PMC5722667

40. Chavali NV, Kryshtal DO, Parikh SS, Wang L, Glazer AM, Blackwell DJ, Kroncke BM, Shoemaker MB, Knollmann BC. Patient-independent human induced pluripotent stem cell model: A new tool for rapid determination of genetic variant pathogenicity in long QT syndrome. Heart Rhythm. 2019;16:1686–1695. PMID:31004778. PMC6935564

41. Wang L, Wada Y, Ballan N, Schmeckpeper J, Huang J, Rau CD, Wang Y, Gepstein L, Knollmann BC. Triiodothyronine and dexamethasone alter potassium channel expression and promote electrophysiological maturation of human-induced pluripotent stem cell-derived cardiomyocytes. J Mol Cell Cardiol. 2021;161:130–138. PMID:34400182.

42. Burridge PW, Matsa E, Shukla P, Lin ZC, Churko JM, Ebert AD, Lan F, Diecke S, Huber B, Mordwinkin NM, Plews JR, Abilez OJ, Cui B, Gold JD, Wu JC. Chemically defined generation of human cardiomyocytes. Nat Methods. 2014;11:855–860. PMID:24930130. PMC4169698

43. Concordet J-P, Haeussler M. CRISPOR: intuitive guide selection for CRISPR/Cas9 genome editing experiments and screens. Nucleic Acids Res. 2018;46:W242–W245. PMID:29762716. PMC6030908

44. Wada Y, Yang T, Shaffer CM, Daniel LL, Glazer AM, Davogustto GE, Lowery BD, Farber-Eger EH, Wells QS, Roden DM. Common Ancestry-Specific Ion Channel Variants Predispose to Drug-Induced Arrhythmias. Circulation. 2022;145:299–308. PMID:34994586. PMC8852297

45. Thomas GP, Gerlach U, Antzelevitch C. HMR 1556, a potent and selective blocker of slowly activating delayed rectifier potassium current. J Cardiovasc Pharmacol. 2003;41:140–147. PMID:12500032.

46. Feaster TK, Cadar AG, Wang L, Williams CH, Chun YW, Hempel JE, Bloodworth N, Merryman WD, Lim CC, Wu JC, Knollmann BC, Hong CC. Matrigel mattress: A method for the generation of single contracting human-induced pluripotent stem cell-derived cardiomyocytes: A method for the generation of single contracting human-induced pluripotent stem cell-derived cardiomyocytes. Circ Res. 2015;117:995–1000. PMID:26429802. PMC4670592

47. Martin M. Cutadapt Removes Adapter Sequences From High-Throughput Sequencing Reads. EMBnetjournal. 2011;17:10–12.

48. Dobin A, Davis CA, Schlesinger F, Drenkow J, Zaleski C, Jha S, Batut P, Chaisson M, Gingeras TR. STAR: ultrafast universal RNA-seq aligner. Bioinformatics. 2013;29:15–21. PMID:23104886. PMC3530905

49. Liao Y, Smyth GK, Shi W. featureCounts: an efficient general purpose program for assigning sequence reads to genomic features. Bioinformatics. 2014;30:923–930. PMID:24227677.

50. Zhao S, Guo Y, Sheng Q, Shyr Y. Advanced heat map and clustering analysis using heatmap3. Biomed Res Int. 2014;2014:986048. PMID:25143956. PMC4124803

51. Love MI, Huber W, Anders S. Moderated estimation of fold change and dispersion for RNA-seq data with DESeq2. Genome Biol. 2014;15:550. PMID:25516281. PMC4302049

52. Wang J, Vasaikar S, Shi Z, Greer M, Zhang B. WebGestalt 2017: a more comprehensive, powerful, flexible and interactive gene set enrichment analysis toolkit. Nucleic Acids Res. 2017;45:W130– W137. PMID:28472511. PMC5570149

53. Subramanian A, Tamayo P, Mootha VK, Mukherjee S, Ebert BL, Gillette MA, Paulovich A, Pomeroy SL, Golub TR, Lander ES, Mesirov JP. Gene set enrichment analysis: a knowledge-based approach for interpreting genome-wide expression profiles. Proc Natl Acad Sci U S A. 2005;102:15545–15550. PMID:16199517. PMC1239896

