## Supplemental methods, tables, and figures for "Increased L-type calcium current causes action potential prolongation in Jervell and Lange-Nielsen syndrome and is a drug target"

**SUPPLEMENTAL MATERIALS**

**Supplemental Methods**

**Supplemental Tables (S1-S5)**

**Supplemental Figures (S1-S9)**

**Supplemental Data (S1-4) are provided in a separate file (.xls)**

### Supplemental Methods

#### *Generation of iPSCs and cardiac differentiation*

Skin biopsy samples (control and JLN1) or peripheral blood mononuclear cells (PBMCs) (JLN2) from patients and a healthy volunteer were obtained for iPSC generation after obtaining written consent under IRB approval (#090544, #9047) in accordance with the Declaration of Helsinki.

Skin fibroblasts and PBMCs were reprogrammed using the Epi5 Episomal iPSC Reprogramming Kit (Invitrogen). Three clones from the control, JLN1, and JLN2 iPSCs line were screened for large chromosomal abnormalities by commercial karyotype analysis (Genetics Associates). One clone from each line displaying a normal karyotype was carried forward for study. Stemness of iPSCs was validated for the presence of stem cell-associated markers via immunofluorescent detection of OCT4 (Cell Signaling, #2750), SSEA4 (DSHB, MC-813-70), SSEA3 (Millipore, MAB4303), and Tra-1-60 (Millipore, MAB4360), as previously reported.[37,38] We have previously reported the molecular and cellular properties, including stemness, of the control iPSC line.[37–41] This line was used as a template for genome-editing to generate an isogenic iPSC line in the present study.

iPSCs were maintained on Matrigel (Corning, #356230) in mTeSR plus basal medium (STEMCELL, #100-0276) and differentiation into iPSC-CMs was accomplished by the monolayer chemical method as previously described.[42] Differentiation media are listed in Supplemental Table S2. In brief, on the day 0 of differentiation, mTeSR plus basal medium was replaced with M1 medium supplemented with CHIR 99021 (STEMCELL, #72054, 6.0 μM). On day 3, medium was replaced with M1 medium supplemented with IWR (Sigma, #I0161, 5 μM). On day 10, M1 medium was replaced with M2 medium. On day 15, iPSC-CMs were purified by enzymatic dissociation. In brief, iPSC-CMs were treated with TrypLE Select Enzyme 1x (Gibco, #12563011) at 37$^{o}$C for 10 mins and harvested in 1xPBS(-). After centrifugation at 250g for 5 mins, iPSC-CMs were resuspended in M2 medium supplemented with 10% FBS and filtered through 100 μm cell strainer (Corning, #431752; Fisher Scientific, #22363549). 0.7-1.5 million purified iPSC-CMs were replated onto matrigel-coated 6-well plate. On the next day, medium was replaced with M3 medium supplemented with Triiodothyronine (Sigma, #T2877, 0.1 μM) and dexamethasone (Cayman, #11015, 1 μM) (M3+TD). On the day 30, M3+TD medium was replaced with M3 medium. iPSC-CMs were studied at days 35-49 after the beginning of differentiation. We have previously reported on electrophysiological and transcriptional maturation of control iPSC-CMs treated with triiodothyronine and dexamethasone with a focus on potassium channels.[41] The control line used here was the same as the one used in the previous report.

***CRISPR/Cas9 genome editing and off-target screening***

Guide RNAs were designed using the CRISPOR online tool (http://crispor.tefor.net/)[43] and ordered as 24-25 bp oligonucleotides. Oligonucleotide sequences used in the study are listed in Supplemental Table S3. Oligonucleotides (100 μmol/L) were annealed and phosphorylated with T4 polynucleotide kinase. Diluted oligoduplex (1:250 in $H_2O$) was ligated into the pSpCas9(BB)-2A-GFP plasmid (a generous gift from Dr. Feng Zhang, Addgene, #43138) using Golden Gate Assembly with BbsI digestion. The correct insert of the guide RNA was verified by Sanger sequencing by a plasmid-specific primer (5'-GGACTATCATATGCTTACCG-3').

The repair template was designed with a target edit including a synonymous variant introduced to disrupt the PAM site. On the CRISPR experimental day, iPSCs at 80% confluency were harvested in mTeSR Plus basal medium. 5.0-7.0 million iPSCs were resuspended in buffer R (Neon Transfection System, Thermo Fisher) with the guide RNA construct (1 – 2 μg) and a repair template (100 pmol) followed by electroporation (at 1200 V, pulse width 20ms, 2 pulses). Electroporated iPSCs were then replated onto matrigel-coated 6-well plate with mTeSR Plus basal medium supplemented with a Y-27632 (Calbiochem, #688000, 10 μM) and 10% FBS. On the next day, medium was replaced with mTeSR Plus basal medium. 2-3 days after the initial editing, cells were harvested in 1 mL of 1xPBS(-) and sorted on the 5-laser FACS Aria for GFP, indicating successful delivery of the guide RNA construct, onto a matrigel-coated 6well-plate with mTeSR Plus basal medium supplemented with CloneR (STEMCELL, #05888) and 1% penicillin/streptomycin. On the day after single cell sorting, the medium was replaced with mTeSR Plus basal medium. 7-10 days after single cell sorting, each iPSC colony was genotyped to identify clones with the desired edit.

***Cell preparation for electrophysiology***

On day 31-45, iPSC-CMs were dissociated by TrypLE Select Enzyme 10x (Gibco, #A1217701) at 37$^{o}$C for 10 mins and harvested in 1xPBS(-). After centrifugation at 250g for 5 mins, iPSC-CMs were resuspended in M3 medium supplemented with 10% FBS and filtered through 100 μm cell strainer (Corning, #431752; Fisher Scientific, #22363549). 25,000-100,000 singularized iPSC-CMs were replated onto 6-well plate containing five glass coverslips (Fisher scientific, #12-545-80P) per well. Glass coverslips were pre-coated with rat tail collagen-I (Corning, # 354236, diluted at 0.5% in 1xPBS(-)). iPSC-CMs were fed on the next day with M3 medium and every day until one day before study.

***Electrophysiology***

Whole-cell and perforated patch-clamp recordings were carried out using an AxoPatch 200B amplifier (Molecular Devices) and the Digidata 1322A data acquisition system.

Action potentials (APs) were recorded using amphotericin B perforated patch-clamp in Tyrode's solution at 35-37$^{o}$C.[44] APs were elicited by injection of a stimulus current (1-2 nA, 1 ms) at 0.5 Hz. Signals were low-pass filtered at 1 kHz and sampled at 10 kHz. We did not exclude recordings that showed non-ventricular waveforms given the potential changes in AP waveforms due to genetic variants. We excluded recordings from analysis when a cell displayed early afterdepolarization(s) that prevented accurate APD measurement at a fixed pacing frequency. Recorded values were not corrected for liquid junction potentials.

Sensitivity to acute exposure to LTCC antagonists was determined by exposure to increasing concentrations of either diltiazem (Sigma) or verapamil (Fluka). During continuous AP recording, the LTCC antagonist was perfused in serial concentrations from 0.1 μM, 1.0 μM, and 10 μM for 2-3 minutes at each concentration. Drug sensitivity was reported as an absolute change in $APD_{90}$ ($\Delta APD_{90}$).

$I_{Ks}$ and $I_{Kr}$ were recorded in whole-cell mode using extracellular Tyrode's solution at 35-37$^{o}$C. For $I_{Ks}$ recording, the holding potential was -80 mV followed by a depolarization pulse (200 ms) to -40 mV, and then 4 sec test steps between +40 to -40 mV in 20 mV decrements. Signals obtained by the square-pulse were low-pass filtered at 1 kHz and sampled at 10 kHz. After break-in, each cell was exposed to 10 μM nifedipine and 0.5 μM E-4031 to fully block L-type calcium current and $I_{Kr}$, respectively, followed by perfusion with the $I_{Ks}$ blocker HMR-1556 (0.5 μM).[45] $I_{Ks}$ was then measured as the HMR-1556 sensitive current obtained by digital subtraction. Steady-state current was measured by averaging the signals obtained between 3.86-3.91 sec after the onset of the depolarizing pulse.

For $I_{Kr}$ recording, the holding potential was -80 mV followed by a depolarization pulse (200 msec) to -40 mV, and then 1 sec test steps between +20 to -40 mV in 10 mV decrements. Low-pass filter and sampling rate were the same as for $I_{Ks}$. $I_{Kr}$ was measured as the E-4031 sensitive current obtained by digital subtraction in the presence of 10 μM nifedipine. Steady-state and peak tail currents were measured by averaging the signals obtained between 910-960 msec and 1030-1040 msec after the onset of the depolarizing pulse, respectively.

The $IC_{50}$ of Ca channel antagonists for $I_{Kr}$ was determined by continuous recording of peak tail current at -40 mV in whole-cell mode.

We did not determine $IC_{50}$ for $I_{Ca\text{-}L}$ due to rundown which prevented measurement of the channel blocking effect at multiple concentrations in the same cell.

$I_{Na}$ recording was carried out at room temperature using a 50 msec depolarizing pulse from -80 to +60 mV in 10 mV increment from a holding potential of -120 mV. Signals obtained by the square-pulse were low-pass filtered at 10 kHz and sampled at 50 kHz.

$I_{NCX}$ was recorded as an acute caffeine-induced inward current (10 mM) at 37$^{o}$C in a $K^+$-free solution at -70 mV continuous voltage clamp.

***Recording solution and chemicals***

The compositions of external and pipette solutions for all patch-clamp experiments are listed in Supplemental Table S4A-B. Patch pipette resistance was 2-5 MΩ (for AP, $I_{Kr}$, $I_{Ks}$, and $I_{NCX}$ recording) or 0.5-1.5 MΩ (for $I_{Na}$ and $I_{Ca\text{-}L}$ recording) when filled by pipette solution. Chemicals used in the study were reconstituted as listed in Supplemental Table S1 and were freshly dissolved in recording solution before perfusion.

***Intracellular Ca measurements in iPSC-CMs***

iPSC-CMs were dissociated and plated on a Matrigel mattress in a Delta TPG Culture Dish (Fisher).[46] After 3-6 days with daily M3 media changes, cells were incubated with a final concentration of 2 µmol/L Fura-2, AM (Molecular Probes Inc, Eugene, OR) in M3 medium for 8 minutes and then washed twice with 1.2 mmol/L $Ca^{2+}$- containing modified Tyrode's solution with 0.25 mmol/L probenecid for 10 minutes each. The composition of modified Tyrode's solution used for Fura-2 loading and washing was (in mM): 134 NaCl, 5.4 KCl, 1.2 $CaCl_2$, 1.0 $MgCl_2$, 10 Glucose, and 10 HEPES (pH adjusted to 7.4 with NaOH). After Fura-2 loading, Ca transients were recorded from cells that elicited a response during 0.5 Hz electrical field stimulation in modified Tyrode's solution with 2 mmol/L $Ca^{2+}$ for 20 seconds using a dual-beam excitation fluorescence photometry setup (IonOptix Corp.). After turning off field stimulation, SR Ca stores were depleted by adding caffeine (10 mM, Sigma) for 5 seconds to estimate total SR Ca content. Ca data files were analyzed using commercially available analysis software (IonWizard, IonOptix, Milton, MA). All experiments were conducted at room temperature.

***Quantitative PCR***

Total RNAs were extracted by RNeasy Plus Mini Kit (QIAGEN) followed by cDNA creation using SuperScript III (Invitrogen). Quantitative PCR was carried out using TaqMan Gene Expression Assays, TaqMan Fast Universal PCR Master Mix (Applied Biosystems), and a CFX-96 Real-Time PCR system (BioRad). Pre-designed probes were used for all genes tested and listed in Supplemental Table S5. All qPCR reactions were performed in triplicate. The normalized values (to the cardiac troponin T [TNNT2]) were calculated using the comparative ΔΔCt method.

***RNA sequencing and analysis***

RNA quality was verified using a Bioanalyzer quality control test and sequenced on an Illumina NovaSeq 6000 instrument using 150 bp paired-end sequencing. Reads were trimmed to remove adapter sequences using Cutadapt v4.5)[47] and aligned to the Gencode GRCh38.p13 genome using STAR (v2.7.11a).[48] Gencode v38 gene annotations were provided to STAR to improve the accuracy of mapping. Quality control on both raw reads and adaptor-trimmed reads was performed using FastQC (v0.12.1) (www.bioinformatics.babraham.ac.uk/projects/fastqc). FeatureCounts (v2.0.6)[49] was used to count the number of mapped reads to each gene. Heatmap3[50] was used for cluster analysis and visualization. Significantly differential expressed genes with absolute fold change ≥ 2 and FDR adjusted p value ≤ 0.05 were detected by DESeq2 (v1.40.2)[51]. Genome Ontology and KEGG pathway over-representation analysis was performed on differentially expressed genes using the WebGestaltR package (v0.4.6)[52]. Gene set enrichment analysis was performed using GSEA package (v4.3.2)[53] on database (v2022.1.Hs).

***Reverse transcription PCR***

Reverse transcription PCR was performed to amplify the targeted region between exon 5 and 8 of *KCNQ1*, using a forward primer (5'-TCCTGAGGATGCTACACGTC-3') and a reverse primer (5'-CTCTGCTTCTGCTGCACCTT-3'), followed by gel electrophoresis in 2% agarose. The band corresponding to the wild-type *KCNQ1* (in the control sample from the control iPSC-CMs) at 376 bp, along with an additional band at ~250 bp, was cut and extracted using the QIAGEN Gel Extraction Kit. Sanger sequencing with a reverse primer identified an exon 6 skipping in JLN2 iPSC-CMs (Supplemental Figure 1).

***Diltiazem challenge in a patient with JLN***

The study was approved by the IRB (#240535) and registered at ClinicalTrials.gov (NCT06534671). After the patient gave written consent, the study was conducted in the outpatient Clinical Research Center (CRC) at Vanderbilt University Medical Center. Baseline vital signs (12-lead ECG, blood pressure) were measured, and a single dose of intravenous diltiazem (21mg/body; 0.25 mg/kg over 2 minutes) was administered. 12-lead ECG and blood pressure measurements were obtained at 2, 5, 7, 10, 15 and 20 minutes. After 20 minutes, the test ended and the patient remained in the CRC, monitored on continuous telemetry for 2 hours.

### Supplemental Tables

**Supplemental Table S1. Drugs used in electrophysiology study.**

| Drug | Stock concentration (mM) | Diluent | Working concentration (µM) | Source |
|---|---|---|---|---|
| HMR-1556 | 0.5 | DMSO | 0.5 | TOCRIS |
| IBMX | 200 | DMSO | 200 | Sigma |
| Forskolin | 10 | DMSO | 10 | Sigma |
| Nifedipine | 50 | DMSO | 10 | Sigma |
| Diltiazem | 10 | $H_2O$ | 0.1 – 10 | Sigma |
| Verapamil | 10 | $H_2O$ | 0.1 – 10 | Fluka |
| E-4031 | 0.5 | $H_2O$ | 0.5 | Cayman |
| Caffeine* | (-) | External solution* | 10 mM | Sigma |
| Amphotericin B** | (-) | Pipette solution** | 200 µg/mL | Sigma |

*Caffeine was freshly dissolved in external solution for $I_{NCX}$ recording.
**Amphotericin B solubilized powder was freshly dissolved in pipette solution for AP recording.
Ion compositions of recording solutions are detailed in Supplemental Table S4A-B.

**Supplemental Table S2. Media and supplementation for cardiac differentiation.**

| | Basal media | Cat# (source) | Supplementation | Cat# (Source) |
|---|---|---|---|---|
| M1 | RPMI1640 | #11875-093 (Gibco) | B27 minus insulin | #2495300 (Gibco) |
| M2 | RPMI no glucose | #11879-020 (Gibco) | | |
| M3 | RPMI1640 | #11875-093 (Gibco) | B27 | #2415366 (Gibco) |
| M3+TD | RPMI1640 | #11875-093 (Gibco) | B27 | #2415366 (Gibco) |
| | | | Triiodothyronine | #T2877 (Sigma) |
| | | | Dexamethasone | #11015 (Cayman) |

**Supplemental Table S3. Oligonucleotides for CRISPR/Cas9 genome editing**

| ***KCNQ1*** **R518X** | |
|---|---|
| Guide RNA top strand | GUGACCUUAAUGGUGGCCCGA |
| Guide RNA bottom strand | UCGGGCCACCAUUAAGGUCAC |
| Donor DNA repair template | GGTGGCCACTCACAATCTCCTCTCCTCTCTCCACTGCAGGCTGCGGGAACATCATCGGGCCACCATTAAGGTCATTTGACGCATGCAGTACTTTGTGGCCAAGAAGAAATTCCAGGTAAGCCCTGTGCTGAGCCTTCCTGCCCTCAGCCTGCCCCTCGCAGC |

**Supplemental Table S4A. Ion compositions of external solutions used in electrophysiology study.**

| Extracellular solution (mM) | | | | | | |
|---|---|---|---|---|---|---|
| | AP | $I_{Kr}$, $I_{Ks}$ | $I_{Ca\text{-}L}$ | $I_{Ba\text{-}L}$ | $I_{Na}$ | $I_{NCX}$ |
| NaCl | 140 | 140 | | | 10 | 135 |
| KCl | 5.4 | 5.4 | | | | |
| CsCl | | | | | 135 | 5 |
| $NaH_2PO_4$ | 0.33 | 0.33 | | | | |
| TEA-Cl | | | 160 | 160 | | |
| $CaCl_2$ | 1.8 | 1.8 | 1.8 | 1.8 | 1 | 1.8 |
| $BaCl_2$ | | | | 1.8 | | |
| $MgCl_2$ | 0.5 | 0.5 | 1 | 1 | 1 | 1 |
| Glucose | 5.5 | 5.5 | 10 | 10 | 10 | 10 |
| HEPES | 5 | 5 | 10 | 10 | 10 | 10 |
| Nifedipine | | 10 µM | | | 10 µM | |
| $NiCl_2$ | | | | | 200 µM | |
| pH | 7.4 (NaOH) | 7.4 (NaOH) | 7.4 (CsOH) | 7.4 ($Ba(OH)_2$) | 7.4 (CsOH) | 7.4 (NaOH) |
| Temperature | 37°C | 37°C | 37°C | 37°C | Room Temperature | 37°C |

AP, action potential.

**Supplemental Table S4B. Ion compositions of pipette solutions used in electrophysiology study.**

| Pipette | solution | (mM) | | | | |
|---|---|---|---|---|---|---|
| | AP | $I_{Kr}$, $I_{Ks}$ | $I_{Ca-L}$ | $I_{Ba-L}$ | $I_{Na-peak}$ | $I_{NCX}$ |
| NaCl | 5 | 5 | 5 | | 5 | 5 |
| KCl | 150 | 150 | | | | |
| CsCl | | | 145 | 145 | 135 | 135 |
| TEA-Cl | | | | | | |
| $CaCl_2$ | | 2 | 2 | | 2 | |
| $BaCl_2$ | | | | 2 | | |
| EGTA | | 5 | 5 | 5 | 5 | |
| HEPES | 5 | 5 | 10 | 10 | 10 | 10 |
| Mg-ATP | | 5 | 5 | 5 | 5 | 5 |
| Amphotericin B | 200 μg/mL | | | | | |
| pH | 7.2 (KOH) | 7.2 (KOH) | 7.2 (CsOH) | 7.2 ($Ba(OH)_2$) | 7.2 (CsOH) | 7.2 (CsOH) |

AP, action potential.

**Supplemental Table S5. Probes for quantitative PCR and siRNAs for gene-specific knockdown.**

| | |
|---|---|
| **Taqman probes for quantitative PCR** | |
| *KCNQ1* | Hs00923522 (Thermo Fisher) |
| *KCNH2* | Hs04234270 (Thermo Fisher) |
| *SCN5A* | Hs00165693 (Thermo Fisher) |
| *KCNE1* | Hs00899754 (Thermo Fisher) |
| *CACNA1C* | Hs00167681 (Thermo Fisher) |
| *CACNB2* | Hs01100744 (Thermo Fisher) |
| *TNNT2* | Hs00943911 (Thermo Fisher) |
| *ATP1A2* | Hs00265131 (Thermo Fisher) |
| *BIN1* | Hs00184913 (Thermo Fisher) |
| *CBARP* | Hs00611235 (Thermo Fisher) |
| *FKBP1B* | Hs00997683 (Thermo Fisher) |
| *TGFB1* | Hs00998133 (Thermo Fisher) |
| *RRAD* | Hs00188163 (Thermo Fisher) |
| siRNAs for gene-specific knockdown | |
| *KCNQ1* | s531183 (Thermo Fisher) |
| *ATP1A2* | 8892 (Thermo Fisher) |
| *BIN1* | s1341 (Thermo Fisher) |
| *CBARP* | s48605 (Thermo Fisher) and s229858 (Thermo Fisher) were combined |
| *FKBP1B* | s5209 (Thermo Fisher) |
| *TGFB1* | 289162 (Thermo Fisher) |
| *RRAD* | s12348 (Thermo Fisher) |

**Supplemental Figures**

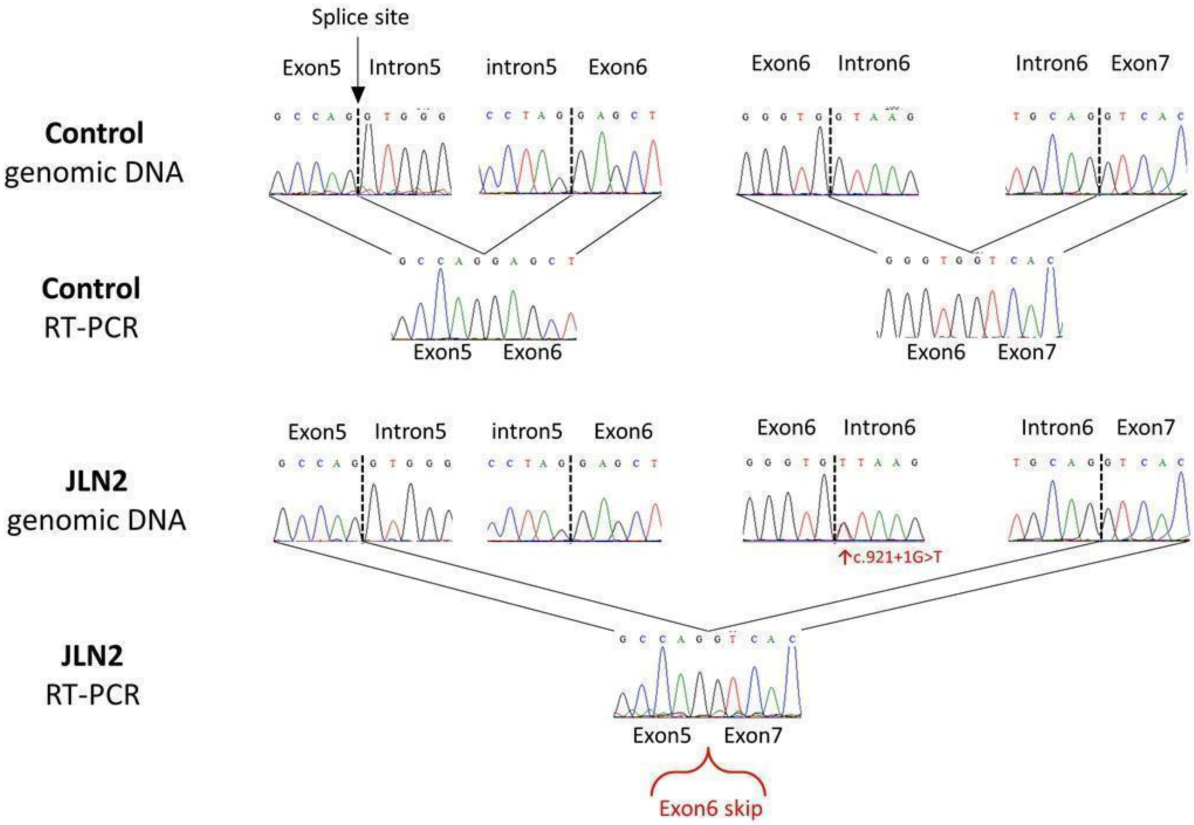


**Figure S1. Sanger sequencing traces of DNA and RNA alignment caused by the splice site variant c.921+1G>T of *KCNQ1* in the patient designated JLN2.**

RT-PCR, reverse transcription PCR.

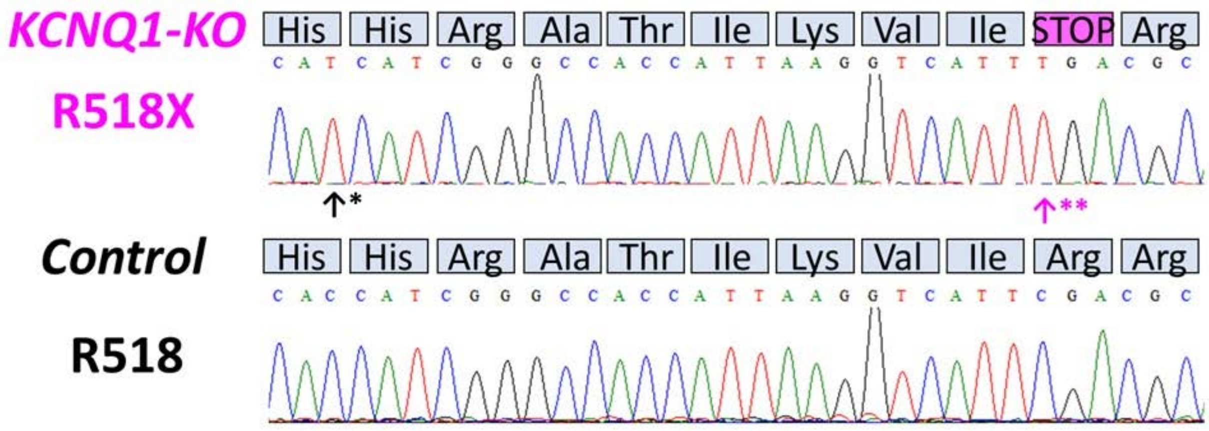


**Figure S2. Sanger sequencing traces of the control and KCNQ1-KO iPSCs.**

The arrow in magenta (**) denotes the R518X variant introduced by genome-editing.

The arrow in black (*) denotes a synonymous variant introduced by genome-editing at the protospacer adjacent motif (PAM) site to prevent re-editing.

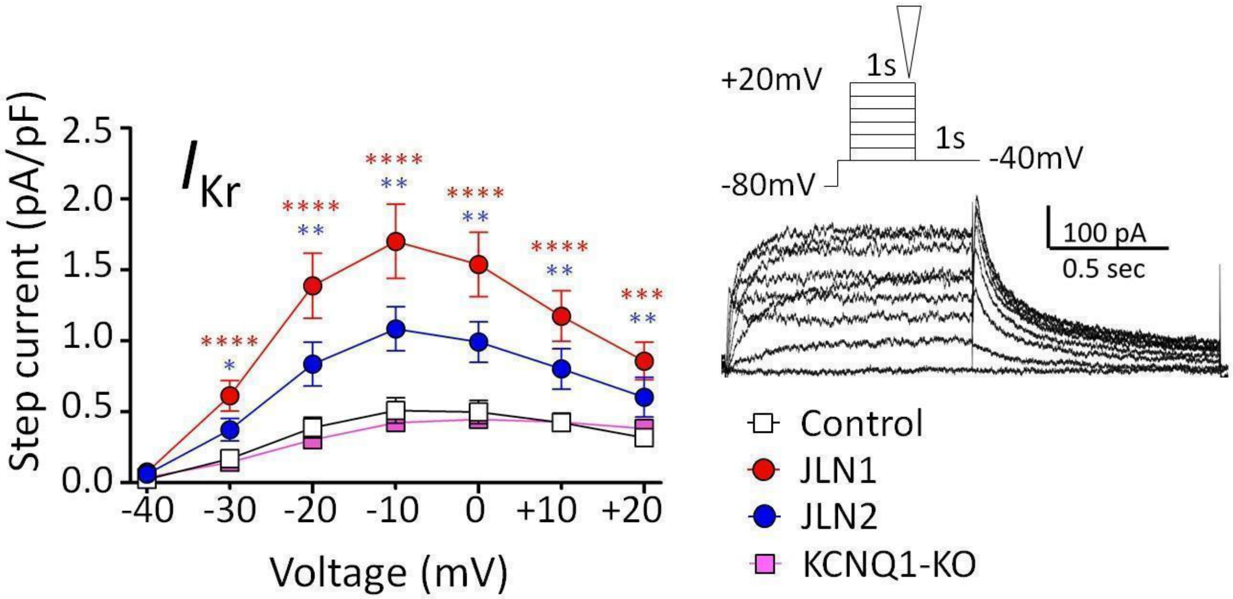


**Figure S3. $I_{Kr}$ in the control, JLN1, JLN2, and KCNQ1-KO iPSC-CMs.**

The *I*-*V* relation for $I_{Kr}$ measured at the end of the depolarizing step as indicated by an inverted triangle in an inset showing representative traces of an E-4031 sensitive current ($I_{Kr}$). n/N=12/4 (JLN1), 9/2 (JLN2), 13/2 (KCNQ1-KO), and 21/9 (control).

Number of cells is expressed as n/N, where n indicates number of recordings and N indicates number of differentiation batches.

*p<0.05, **p<0.01, ***p<0.001, ****p<0.0001 against the control cells by Kruskal-Wallis test. Red and blue asterisks denote statistics of JLN1 and JLN2 cells against control cells, respectively.

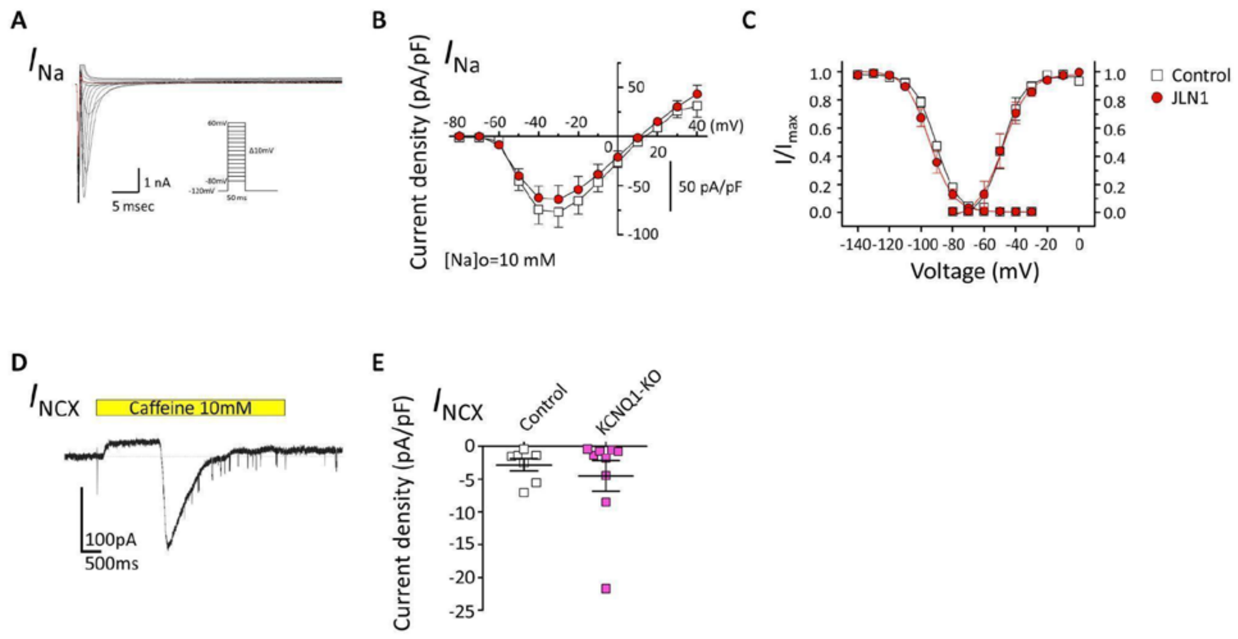


**Figure S4. Sodium current and sodium-calcium exchanger current in iPSC-CMs with genetic ablation of *KCNQ1*.**

***A.*** Representative traces of sodium current ($I_{Na}$) in the control cell. The inset shows voltage protocol. ***B.*** The *I-V* relation for $I_{Na}$ measured at the peak inward current during the depolarizing step. Control cells (n/N=7/1) in the open squares. JLN1 cells (n/N=6/1) in the red circles. ***C.*** Voltage dependence of activation and inactivation of $I_{Na}$ for the control (open squares) and JLN1 (red circles) cells. ***D.*** Representative trace of sodium-calcium exchanger current ($I_{NCX}$) elicited by 10 mM caffeine as shown by a yellow bar. ***E.*** Current densities of $I_{NCX}$ for the control (open squares, n/N=7/1) and KCNQ1-KO (magenta squares, n/N=9/2) cells.

Number of cells is expressed as n/N, where n indicates number of recordings and N indicates number of differentiation batches.

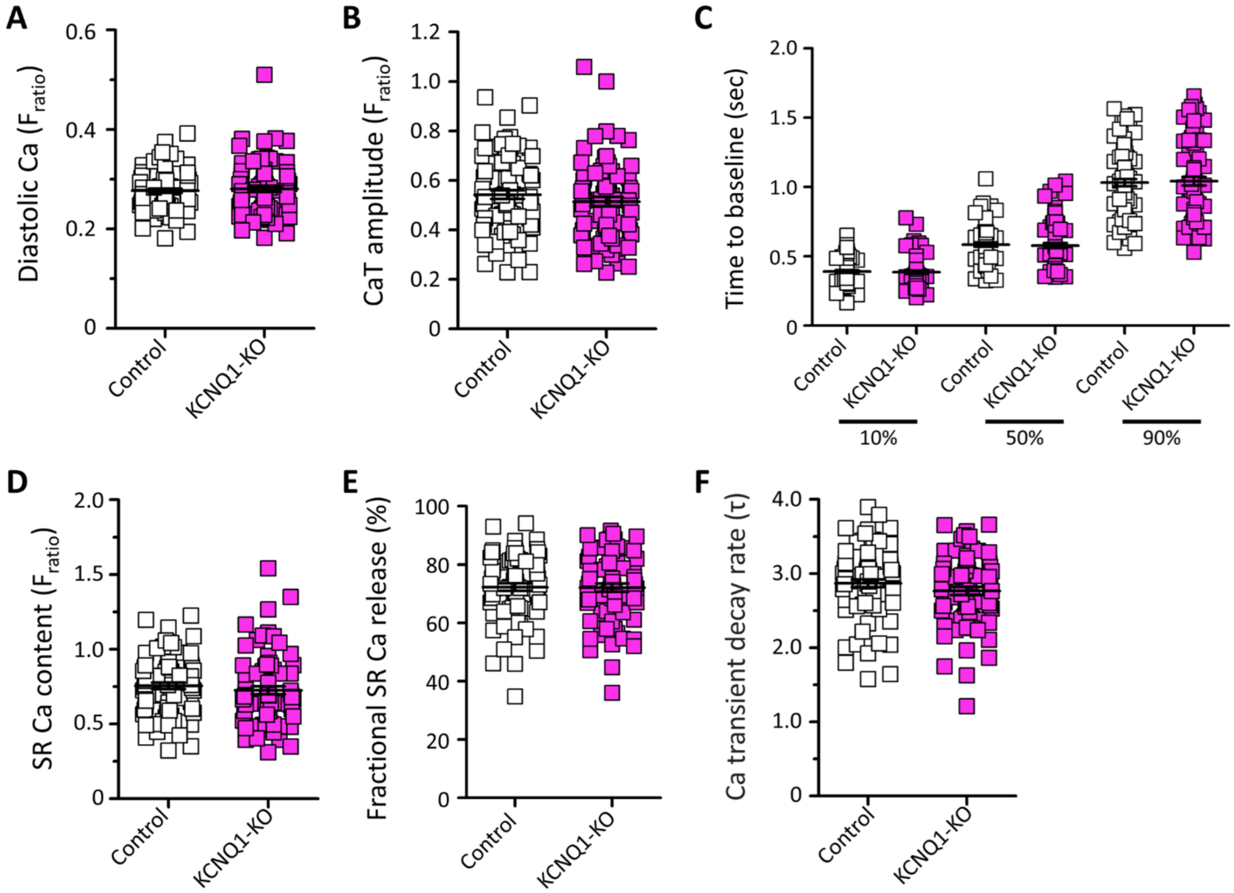


**Figure S5. Calcium transient measurements in iPSC-CMs with genetic ablation of *KCNQ1*.** Control cells in open squares (n/N=82/1), KCNQ1-KO cells in magenta squares (n/N=77/1). There were no significant differences between control and KCNQ1-KO cells in each measurement. Number of cells is expressed as n/N, where n indicates number of recordings and N indicates number of differentiation batches.

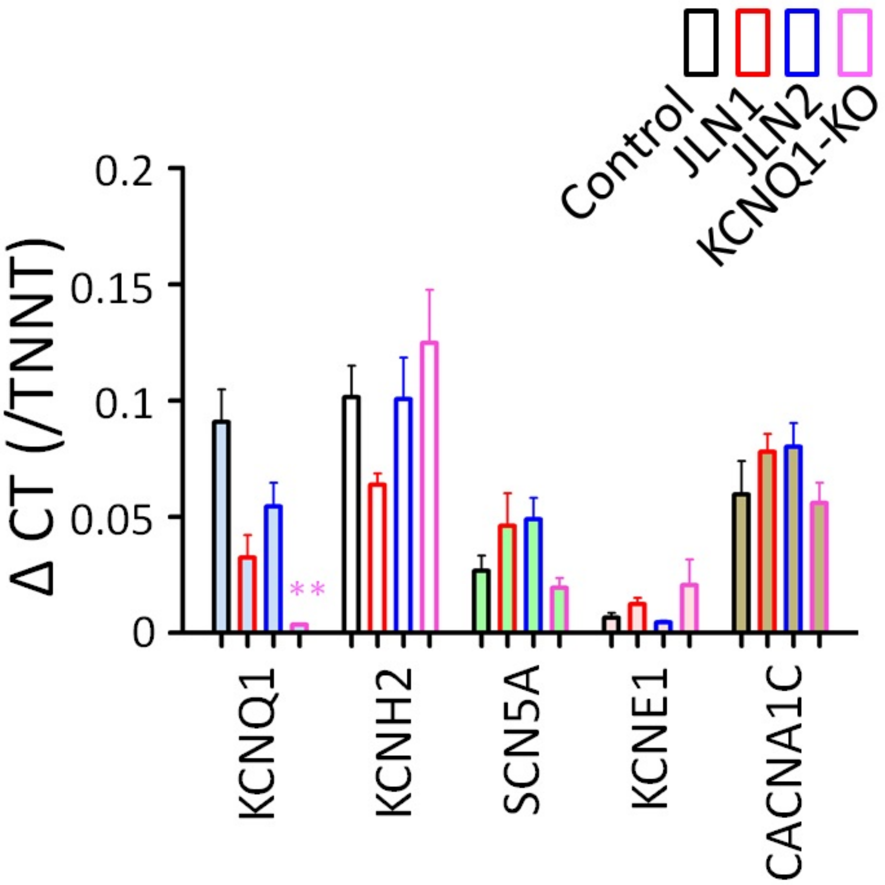


**Figure S6. Expression of major cardiac ion channel genes in the control and JLN cells.**

Quantitative PCR assessed gene expression in the control, JLN1, JLN2, and KCNQ1-KO iPSC-CMs. n=3-6 replicates per gene.

**p<0.01 against the control cells by Kruskal-Wallis test.

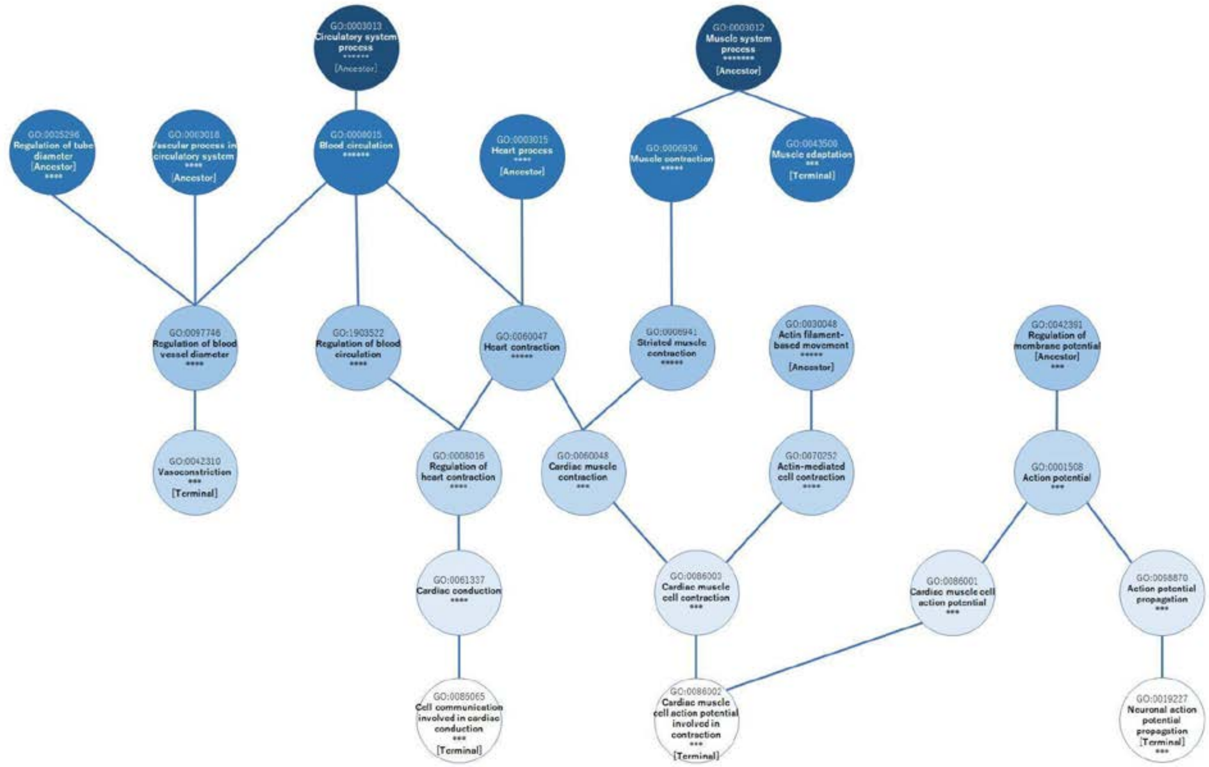


**Figure S7. Gene ontology analysis of genes differentially expressed between control and isogenic KCNQ1-KO lines identified pathways associated with cardiac conduction and contraction.**

GO accession numbers and gene names involved in the pathway are listed in Supplemental Data S2.

***p<0.001, ****p<1.0E-4, *****p<1.0E-5, ******p<1.0E-6.

Ancestor/Terminal denotes the most upstream/downstream pathway in the biological axis, respectively.

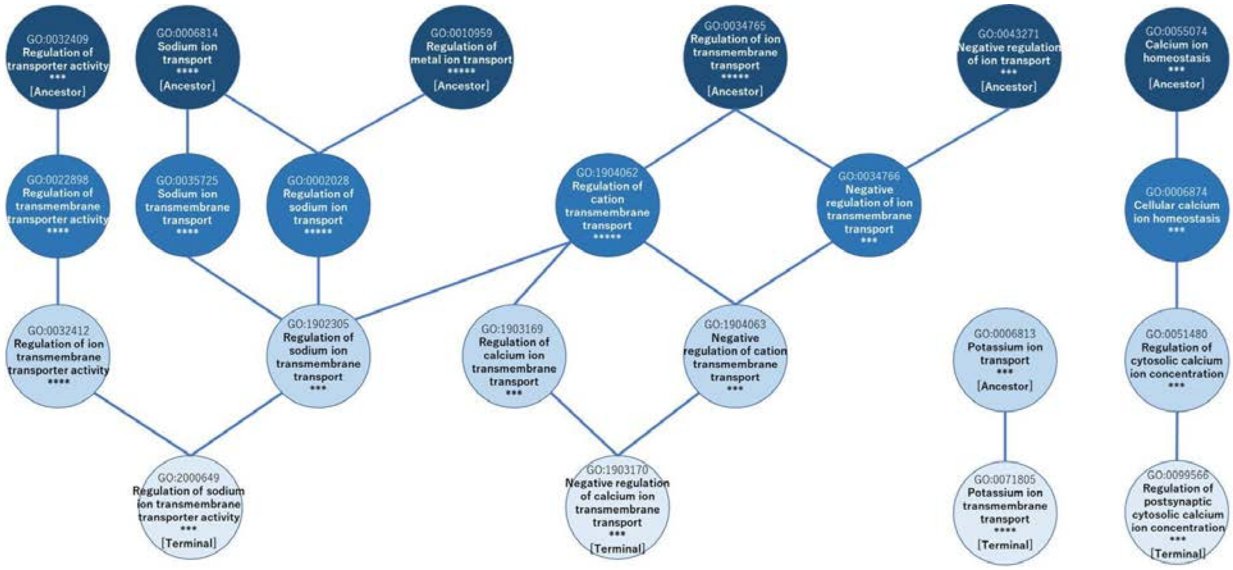


**Figure S8. Gene ontology analysis of genes differentially expressed between control and isogenic KCNQ1-KO lines identified pathways associated with sodium, calcium, and potassium ion transport regulation.**

GO accession numbers and gene names involved in the pathway are listed in Supplemental Data S2.

***p<0.001, ****p<1.0E-4, *****p<1.0E-5.

Ancestor/Terminal denotes the most upstream/downstream pathway in the biological axis, respectively.

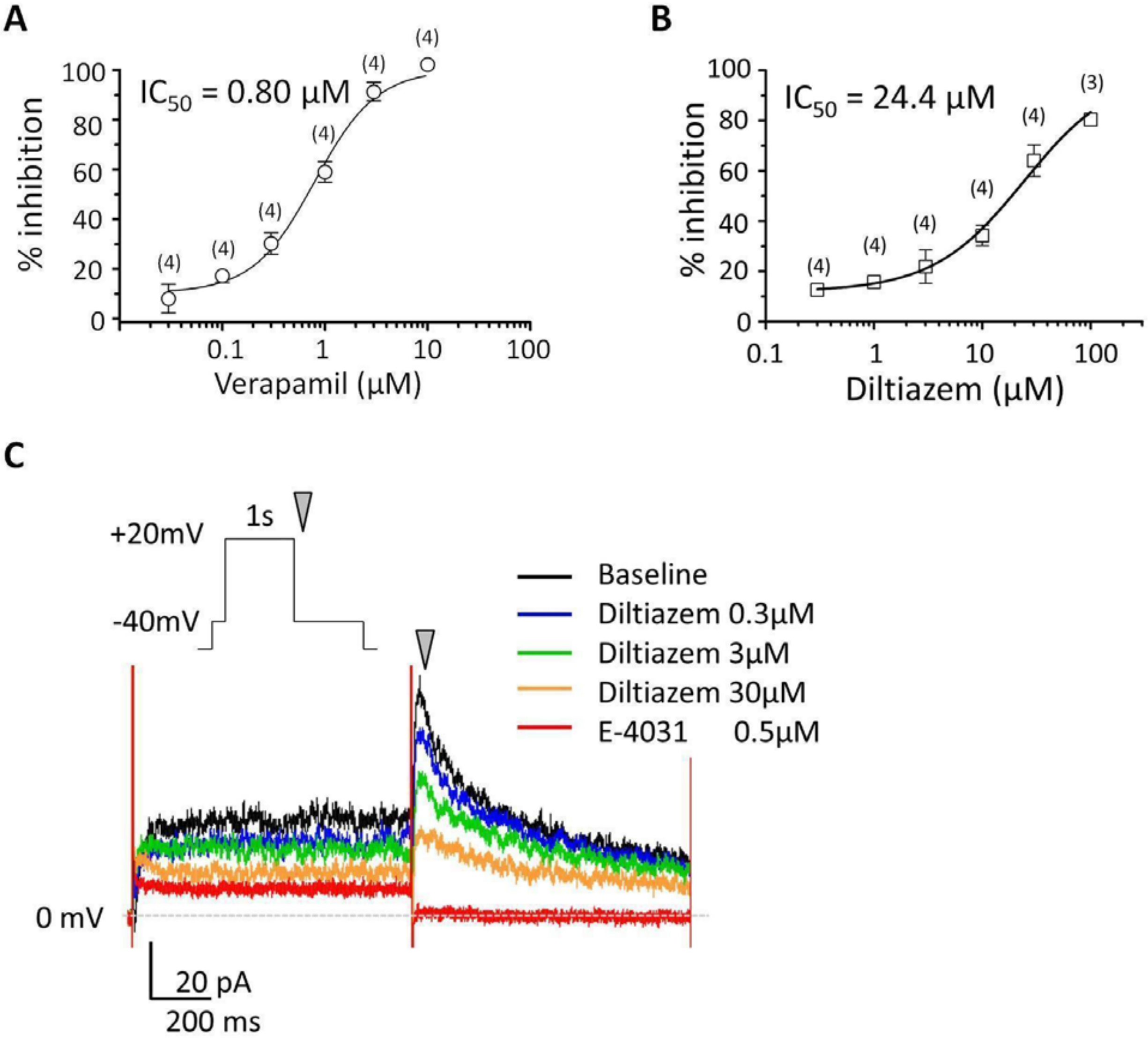


**Figure S9. Effects of Ca channel antagonists on $I_{Kr}$ in the control iPSC-CMs.**

**A-B.** Inhibition as a function increasing drug concentration allowed calculation of a half-maximal inhibitory concentration (IC$_{50}$) of verapamil (***A)*** and diltiazem (***B***) in the control iPSC-CMs. Numbers on each plot indicate the number of recordings tested. **C.** Representative raw traces of $I_{Kr}$ measurement for IC$_{50}$ determination. $I_{Kr}$ was continuously recorded in serially increasing concentrations of diltiazem (0.3 - 30 µM for brevity). The inset indicates the voltage protocol. The inverted gray triangle in the inset corresponds to the one on the traces, indicating where the tail current amplitude was measured to determine the inhibitory effect.
